# A century of weekly notifiable disease incidence data by province in Canada

**DOI:** 10.1101/2024.12.20.24319425

**Authors:** David J. D. Earn, Gabrielle MacKinnon, Samara Manzin, Michael Roswell, Steve Cygu, Chyunfung Shi, Benjamin M. Bolker, Jonathan Dushoff, Steven C. Walker

## Abstract

Canadian notifiable disease surveillance programmes have recorded communicable disease incidence data, dating back to the late 19th century. A Public Health Agency of Canada web-portal provides summaries of these data from 1924–2023, but lacks details on how incidence varies seasonally and geographically among provinces. The sub-annual (weekly, monthly, quarterly) and sub-national (provincial, territorial) data required to study such patterns appear in government documents, but are only available in typewritten or handwritten hard copies. We digitized and collated these sources to make sub-annual and sub-national Canadian disease incidence data conveniently available for researchers. We manually transcribed hard copies into digital spreadsheets resembling the originals, enabling accurate transcription through easier cross-checking. We supplemented these historical data sources with more recent digital spreadsheets obtained directly from two provincial agencies. We standardized and combined these spreadsheets into consistent, machine-readable CSV files containing 1,631,380 incidence values from 1903–2021. Because multiple publications and agencies reproduced case counts from the same surveillance system, and many publications reported the same cases at multiple levels of aggregation, many cases were counted more than once among these 1,631,380 incidence values. We reconciled overlapping counts to produce a dataset containing 934,010 unique incidence values at sub-national and sub-annual scales (829,689 weekly; 567 2-weekly; 82,267 monthly; 20,967 quarterly; 520 3-quarterly) covering 139 diseases stratified by province or territory. We illustrate the value of these sub-annual and sub-national data using two examples: synchronized annual cycles of poliomyelitis across provinces and spatially heterogeneous resurgence of whooping cough in the 1990s. Canada’s infectious disease surveillance has produced a detailed record of sub-annual and sub-national disease incidence data that remains largely unexplored. This record is now available as the Canadian Notifiable Disease Incidence Dataset (CANDID), hosted on a publicly accessible website along with code to reproduce it, and scans of the original sources.

## Introduction

Learning from data on past communicable disease outbreaks and recurrent epidemics is an important component of public health planning [1–5]. The year 2024 marked the 100th anniversary since the Canadian federal government began collecting such data through notifiable disease surveillance programmes [6–10]. Several provinces conducted surveillance before 1924 going back to the late 19th century, although here we focus on the period from 1903 to 2021. The Public Health Agency of Canada (PHAC) provides summaries of these data as annual, national totals since 1924 through an online portal (https://diseases.canada.ca/notifiable). These coarsely aggregated data are useful for understanding broad trends, but provide no information on seasonal patterns of incidence within years or on spatial patterns across provinces. For example, annual data cannot be used to estimate the timing and shape of an outbreak curve, and national data cannot be used to assess whether provinces displayed different patterns of spread in the vaccine era.

The Canadian notification system has at times collected more informative weekly, monthly, and quarterly incidence data, stratified by disease and province/territory, but these data are not available through the portal. We use the term sub-annual to refer to surveillance data reported at a finer temporal resolution than one year (e.g., weekly, monthly, or quarterly), as opposed to annual data aggregated over the full year. Likewise, we use the term sub-national to refer to data reported at a finer spatial resolution than the national level (e.g., by province or territory). These higher-resolution data enable investigation of variation within years and across regions, and can be aggregated to coarser temporal or spatial resolutions when broader comparisons are needed. The reverse, however, is not possible, making higher-resolution data inherently more versatile.

It has generally been prohibitively challenging for researchers to find and access these sub-annual and sub-national data. Still, we show here that much of this information exists in government publications (either as hard copy reference material or online), in data and documents obtained directly from government agencies, and in provincial publications, some of which predate the establishment of the federal system in 1924. This information appeared under a variety of evolving titles often issued by Statistics Canada (and its predecessor the Dominion Bureau of Statistics), Health Canada, and provincial and territorial health departments. The diversity of publication formats and naming conventions has made systematic retrieval challenging. To address this, we have documented the available resources in this literature (see Section A in S1 Appendix) and digitized the data they contain using open-source tools that we developed. Here, we introduce CANDID (Canadian Disease Incidence Dataset), a curated dataset that integrates and cleans these disparate data sources to create a comprehensive and accessible digital record of sub-annual (weekly, monthly, quarterly) and sub-national (provincial, territorial) Canadian notifiable disease incidence data–including substantial amounts of data that were publicly available in principle but almost entirely overlooked. Our objective is to make these sub-annual and sub-national disease incidence data, which have been difficult to obtain, conveniently available for public health and research use.

With this paper we announce the existence of CANDID on a publicly available web site (Section C in S1 Appendix) and illustrate its value. Two examples illustrate the advantages of the sub-annual and sub-national data provided by CANDID. First, we describe how poliomyelitis incidence was strongly and consistently seasonal from 1933 to 1963, and that the yearly peaks were synchronous across provinces. This pronounced synchrony contrasts with patterns in the United States [11], but can be interpreted within the context of a broader latitudinal gradient in the timing of seasonal epidemic peaks across North America. Second, the well-studied apparent resurgence of whooping cough in the 1990s showed significant regional variation. While the territories, prairie provinces, and Qúebec experienced clear increases in incidence, this pattern was not evident in British Columbia, Ontario, and the Atlantic region. We use a simple graphical approach in these examples, illustrating how these data can inform fundamental questions in epidemiology. Formal statistical analyses that dig deeper into specific questions will follow in subsequent publications. The Canadian Disease Incidence Dataset (CANDID) has the potential to drive many studies by the broader public health research community.

## Materials and methods

### Data sources

We began searching for Canadian historical infectious disease notification data in 2000. We focused on sub-annual and sub-national data collected through the surveillance programs described in Section A in S1 Appendix, particularly incidence over entire provinces and territories, since finer spatial resolution data were rare. We compiled provincial and territorial population data [12–14] to compute comparable incidence rates per 100,000. We acquired data in any of three formats: (1) paper hard copies, (2) digitally produced PDF files, or (3) spreadsheets (including CSV files). We scanned all of the hard copies. We found that Optical Character Recognition (OCR) was unable to convert scans into digital spreadsheets with sufficient accuracy, so we entered the data manually (see the Data entry section). We used PDF extraction tools [15] to avoid manual entry of digitally-produced PDF pages.

### Data entry

We manually entered the information in scans into replica Excel spreadsheets (i.e., digital spreadsheets in which the layout of each spreadsheet matches the original), to facilitate comparing reproductions with their sources (Fig 1). The Ontario Ministry of Health data were entered before our comprehensive effort and were only partially entered as replicas (they are the sole exception). Reading scans and interpreting handwritten sources (Fig 2) was often a slow, and occasionally error-prone, process. Where available in these sources, we also entered reported annual and national data to support later validation against marginal totals (see the Quality control section). When we encountered unclear numbers in the source material, we recorded our initial guesses using a predetermined format described in Section E in S1 Appendix, so that they could be processed systematically. These guesses were revisited and refined as needed when validation procedures, which compared sums of incidence values with marginal totals reported in both CANDID sources and the PHAC portal [10], revealed discrepancies. Scripted data preparation pipelines (see the Data preparation section) facilitated updates throughout this process. We provide details on our data entry process in Section B in S1 Appendix.

**Fig 1.**
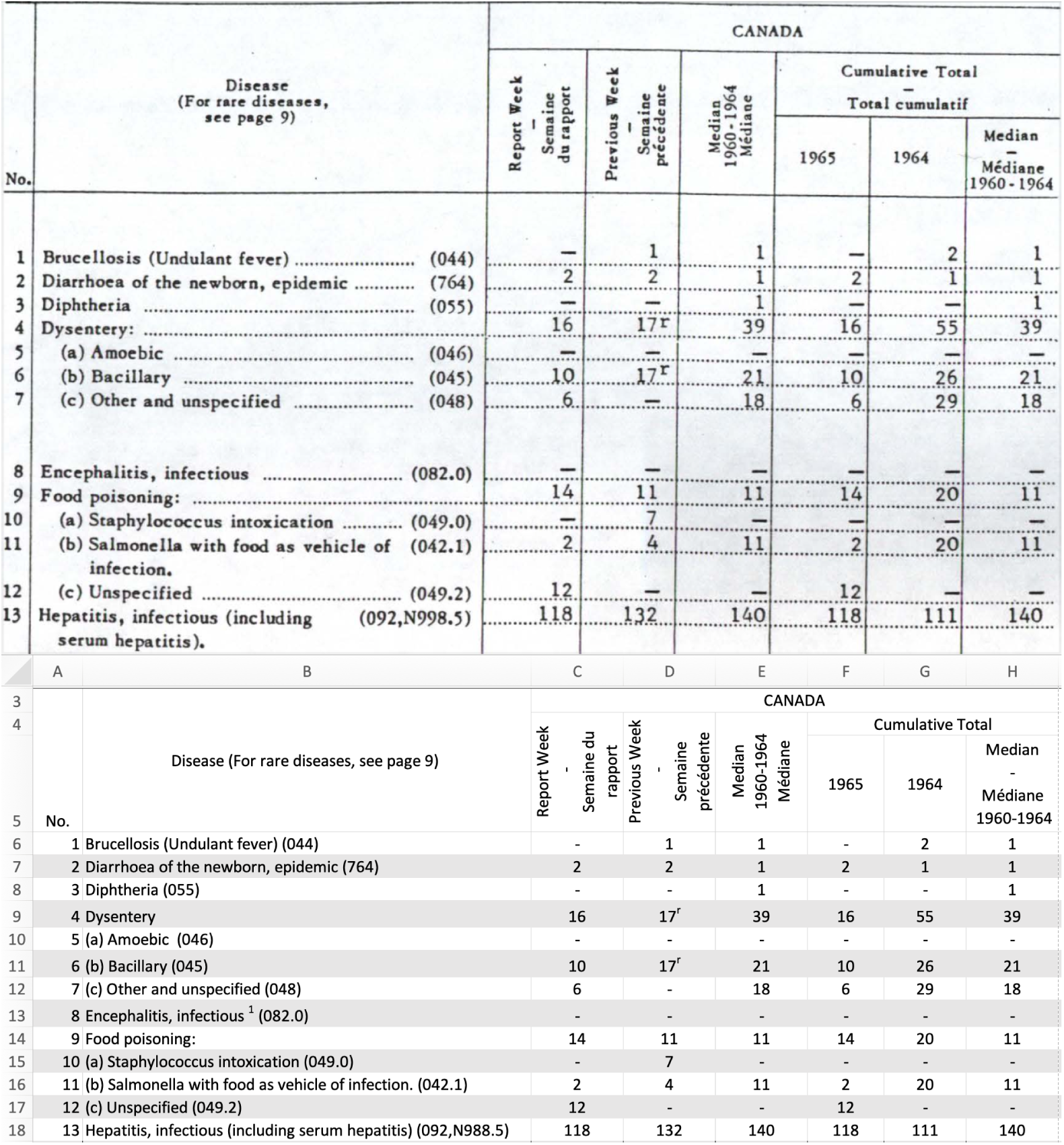
Example of a typewritten source document prepared using a typewriter. The top panel shows a scan of part of this source document, and the bottom panel shows our replica in Microsoft Excel of this same part.

**Fig 2.**
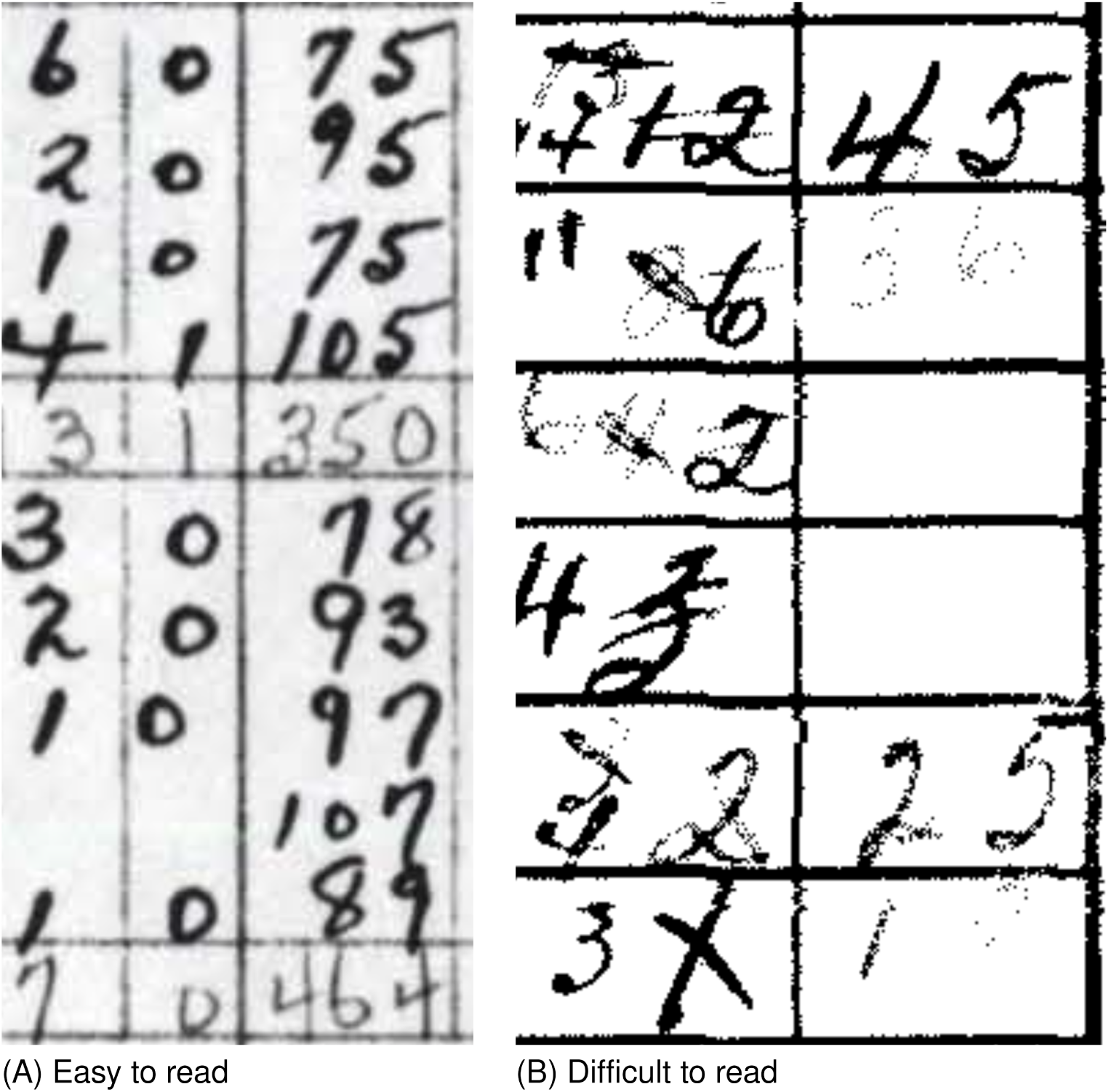
Examples of handwritten data. Most handwritten hard copies, such as the 1939 erysipelas and gonococcal data from the Ontario Ministry of Health (A), are easy to read. Others, like the 1955 poliomyelitis data from Statistics Canada (B), are difficult to read, posing challenges for digitization.

### Data preparation

We developed open-source [16] pipelines that convert replica spreadsheets (Fig 1) into CSV files, using common variables to combine data from different sources (details in Sections D-G in S1 Appendix). The focal variable is the number of new cases of a specific disease reported in a specific location over a specific time period. CANDID consists of three CSV files, ranging from a minimally processed file offering maximum flexibility in data preparation to a heavily processed file prioritizing convenience; each CSV file corresponds to a stage in the data processing pipeline (Fig A in S1 Appendix).

The unharmonized file preserves the raw, digitized data, giving researchers the freedom to apply their own data preparation methods. Descriptors in the unharmonized file use original names (e.g., “infantile paralysis” for poliomyelitis before 1924) to minimize historical information loss [17]. The harmonized file removes low quality data, aggregates some municipal data to provincial levels, and adds harmonized location and disease descriptors that simplify the combination of data from different sources (e.g., poliomyelitis whenever infantile paralysis is reported). Harmonized CSV files are convenient for querying diseases, provinces, and time periods, but they cannot be used directly for analysis due to overlapping incidence values. In data science terms, the harmonized data are not normalized [18, 19]. For example, the harmonized data include total polio incidence alongside separate values for polio with and without paralysis (see Section G in S1 Appendix for more examples). Such overlapping data are useful for quality control (which we explain in the Quality control section), but data analysis requires removing overlaps to prevent double-counting cases. The normalized file does not contain overlapping incidence values, enabling aggregation without double-counting. When deciding which overlapping records to remove, we generally retain the finest resolution. Continuing our example, we would remove total polio incidence and retain separate records for polio with and without paralysis (see Section G in S1 Appendix for details on removal criteria). For convenience when computing incidence rates, we joined provincial population sizes to the normalized data. All figures in this paper are based on the normalized file. Researchers who wish to make different harmonization or normalization choices can use the upstream files (see Section G in S1 Appendix for details).

### Data provenance

Each record can be traced back to the relevant original scan, replica spreadsheet, and/or script used to produce it, using information in the CSV files (Section H in S1 Appendix). We follow research data management practices by distributing DataCite [20] (version 4.3) metadata with each dataset in the archive. These metadata will make it easier to deposit future versions of CANDID into a research data repository, which we plan to do.

### Quality control

We compared sums of incidence values with marginal totals reported both in CANDID sources and in the PHAC portal [10]. Discrepancies suggest possible data-entry or scripting errors. We investigated discrepancies and fixed those that appeared to be due to digitization error. These investigations were simplified using our open data provenance tools (see the Data provenance section) and digitized data source replicas (Fig 1). We provide full detail on quality control in Section I in S1 Appendix.

## Results

As of October 2025 CANDID is based on 12 sources (Table 1). Section C in S1 Appendix provides information on how to access the resulting 1,631,380 unharmonized, 1,186,778 harmonized, and 934,010 normalized incidence values for the provinces and territories of Canada (Fig 3).

**Fig 3.**
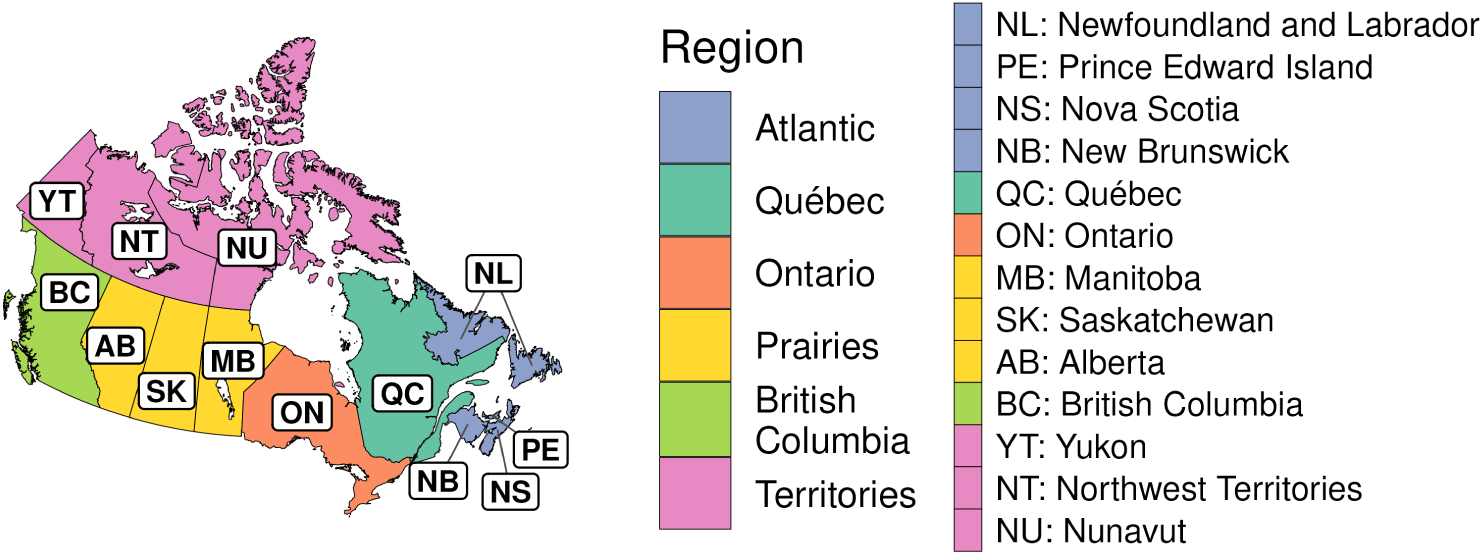
Map of Canada with provinces and territories labelled. The map is used to show the order of provinces and territories in Fig 4, Fig 5, Fig 6, and the order of provinces in the poliomyelitis example (see the Results section). The map also illustrates the grouping of provinces and territories into regions that are used in the whooping cough example (see the Results section). This map was prepared using the rnaturalearth R package [21], which utilized the following public domain shapefile: https://github.com/nvkelso/natural-earth-vector/blob/ e08a35c801ac729401e2de9f5eef206031e6a284/10m_cultural/ne_10m_admin_1_ states_provinces.shp.

**Table 1.** Data sources. The Frequency column gives the shortest period over which incidence counts were reported for all diseases and locations in the source (if not all disease-location combinations have the shortest period, multiple frequencies are given). Sources that include handwritten data are indicated in the Received As column. Details on these sources are provided in Section A in S1 Appendix.

| Years | Provinces | Frequency | Organization | Received As |
| --- | --- | --- | --- | --- |
| 1903-1947 | Ontario | monthly | Ontario Ministry of Health | Hard copy |
| 1910, 1921-1927 | Saskatchewan | monthly | Saskatchewan Department of Public Health | Hard copy |
| 1915-1925 | Québec | monthly | Québec Ministry of Health and Social Services | Hard copy |
| 1924-1955 | All | weekly, monthly | Statistics Canada | Hard copy (with handwriting) |
| 1939-1989 | Ontario | weekly | Ontario Ministry of Health | Hard copy (with handwriting) |
| 1956-1978 | All | weekly | Statistics Canada | Hard copy |
| 1979-1989 | All | 4-weekly | Statistics Canada | Hard copy |
| 1990-2001 | All | monthly, quarterly | Health Canada | Hard copy |
| 1990-2021 | Ontario | weekly | Public Health Ontario | Spreadsheet |
| 2001-2006 | All | quarterly | Canada Communicable Disease Report | PDF |
| 2004-2017 | Manitoba | monthly | Public Health Manitoba | PDF |
| 2004-2019 | Alberta | weekly | Alberta Health | Spreadsheet |

Fig 4, Fig 5, and Fig 6 list the 139 diseases that appear in the normalized dataset, and highlight the time periods in which weekly, monthly, or quarterly incidence data were found in each province or territory. Within each disease, vertical placement indicates data availability for a particular province or territory, ordered approximately clockwise as indicated on the map in Fig 3. The case numbers for many of these diseases are aggregated from 315 “sub-diseases” (Section F in S1 Appendix), harmonizing 929 unique historical name variants. The stratification of each disease into sub-diseases varied across sources (details in Section G in S1 Appendix).

**Fig 4.**
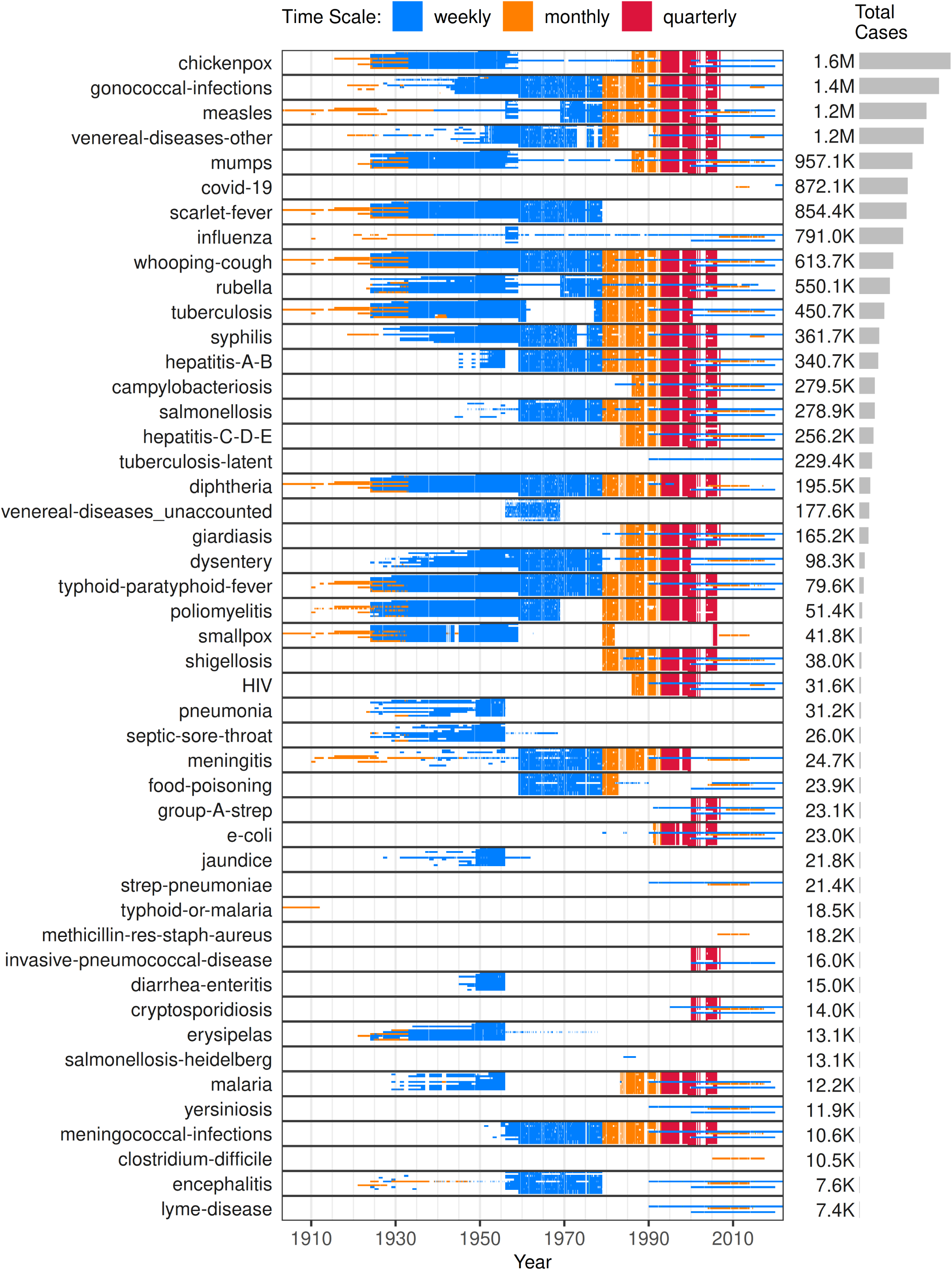
Data availability for highly-reported diseases (top 47 by total reported cases). Moderately- and rarely-reported diseases appear in Fig 5 and Fig 6. The diseases are ranked by the total number of cases (right panels), summed over all provinces for which data were obtained. The diseases are ordered with the largest number of cases at the top. Each incidence value, including zeros, is shown as a tiny coloured rectangle. The y-axis labels identify the disease, while the rectangle’s length along the x-axis represents the temporal extent. Colours indicate reporting frequency: weekly (blue), monthly (orange), and quarterly (red). Two-weekly data are shown in green but are not included in the legend because they are too few to see without zooming. Three-quarterly data are omitted, as they are few and span most of a year (April-December 1997), providing little additional information on within-year variation. White spaces represent missing data (see Section E in S1 Appendix for details on the varied reasons for missing data). The vertical position within each disease denotes the province/territory, arranged roughly clockwise (east-to-west for provinces and then west-to-east for territories): NL-NS-PE-NB-QC-ON-MB-SK-AB-BC-YT-NT-NU (Fig 3). Thin horizontal patterns arise because provinces and territories differ in data availability and reporting frequency: when data are available for only some provinces, or when provinces and territories report at different time scales, the resulting variation appears as horizontal features due to the fixed top-to-bottom ordering of provinces and territories within each disease.

**Fig 5.**
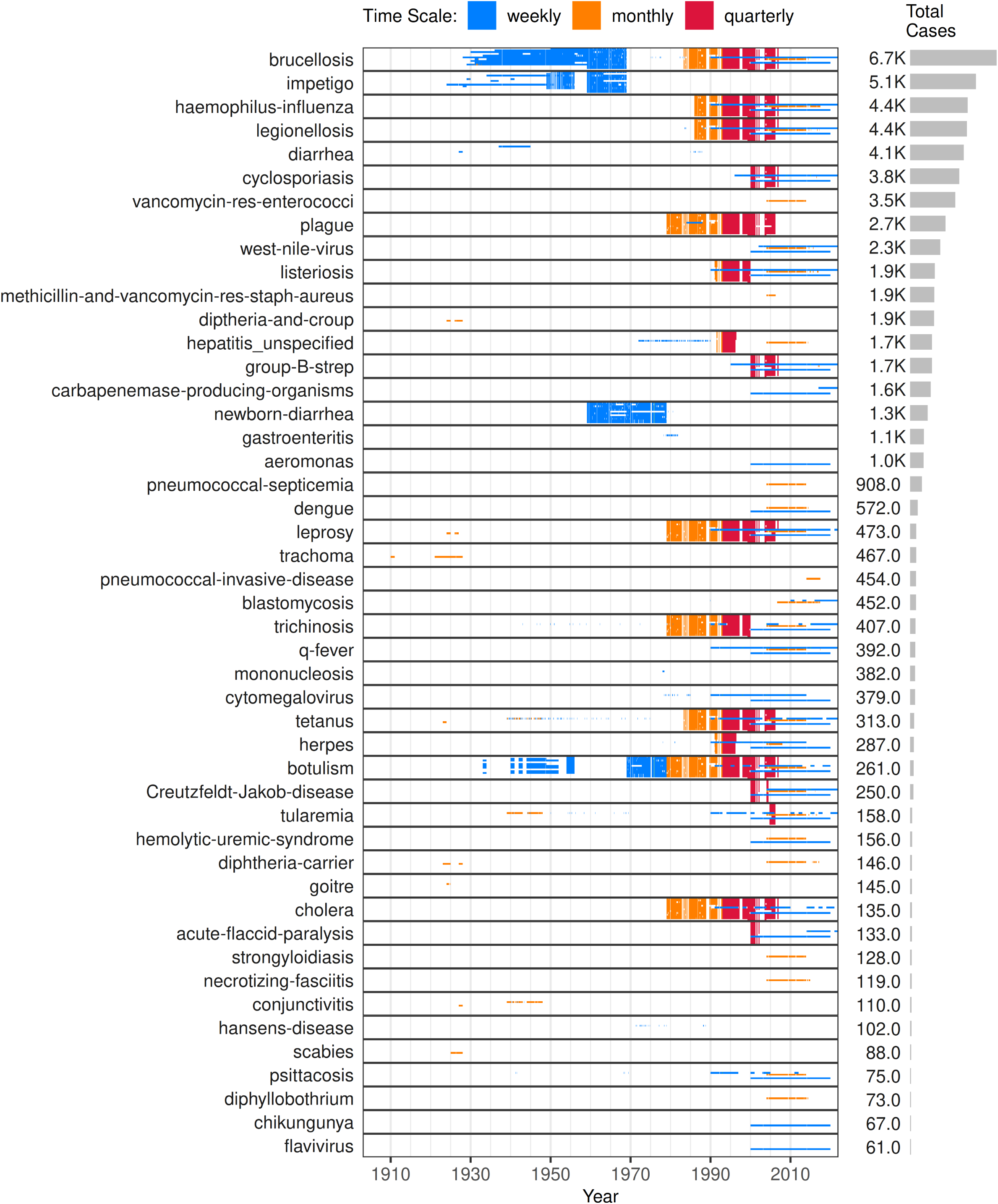
Data availability for moderately-reported diseases (middle 47 by total reported cases). Highly- and rarely-reported diseases appear in Fig 4 and Fig 6. Please see the caption for Fig 4 for a full description of all of these plots.

**Fig 6.**
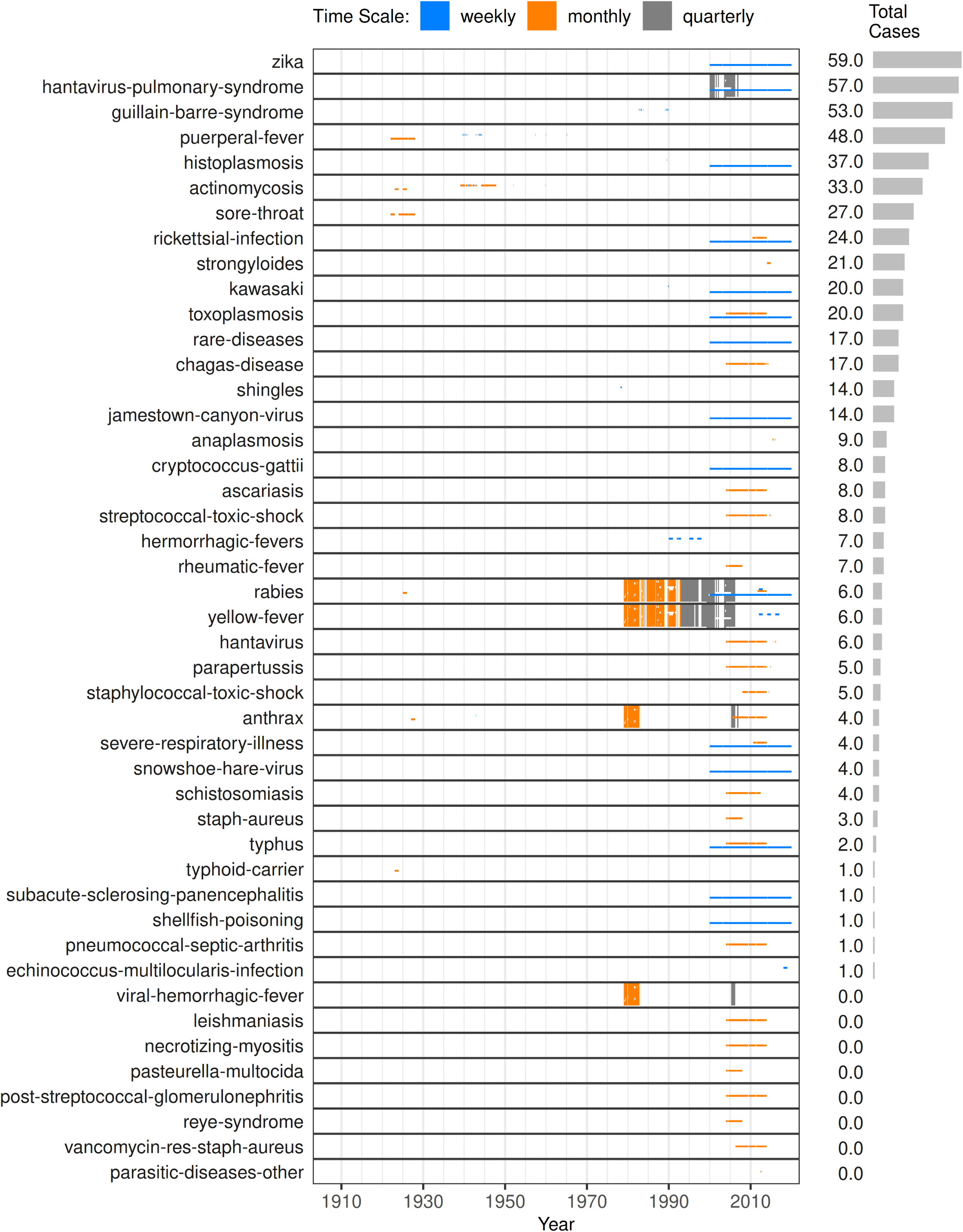
Data availability for rarely reported diseases (bottom 45 by total reported cases). Highly- and moderately-reported diseases appear in Fig 4 and Fig 5. Please see the caption for Fig 4 for a full description of all of these plots.

### Examples

#### Poliomyelitis cases peaked at the same time each year across all provinces

Poliomyelitis incidence was strongly seasonal (upper panel, Fig 7), with national peaks consistently occurring between week 31 and 40 each year from 1933 to 1963, after which the cycles disappeared. These annual cycles were synchronous across provinces, with peak incidence occurring around the same time in each (provincial panels, Fig 7). All national peaks fell between August and October, and most provincial peaks followed this pattern, with only a few outlying province-year combinations (Fig I in S1 Appendix). Identifying such spatial synchrony requires incidence data at sub-annual and sub-national scales.

**Fig 7.**
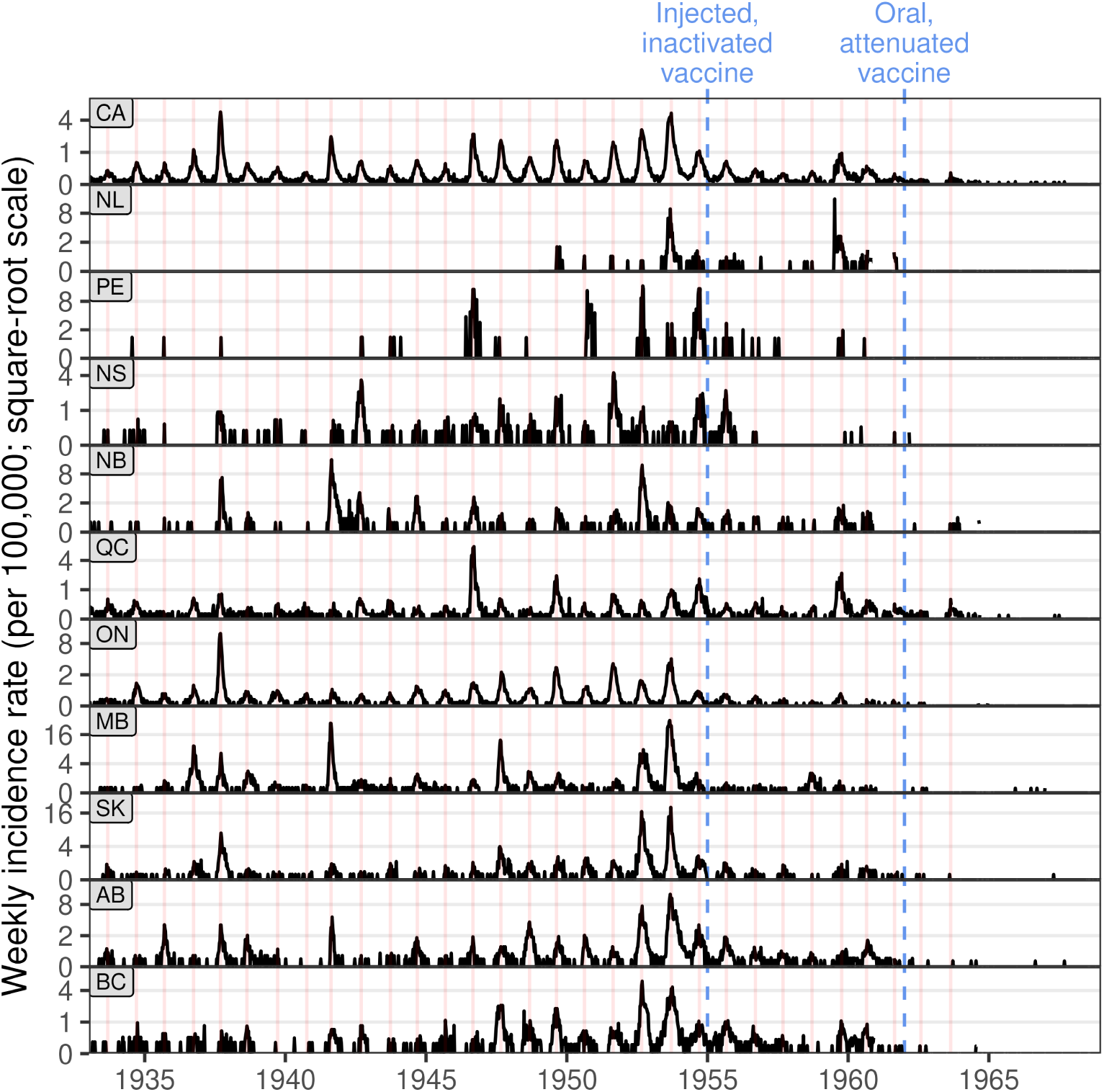
Weekly poliomyelitis incidence from 1933 to 1968 (square root scale). The vertical lines do not indicate the start of each year but mark the week of peak national incidence (top panel) in years with more than 20 cases. Provincial peaks closely align with these national peaks, indicating strong spatial synchrony. This pattern could not have been detected with annual or national data. The introduction of two important vaccination programmes are shown as blue vertical dashed lines. We plot incidence rates per 100,000 to make incidence comparable across provinces and territories, and with other studies. Methods for producing this plot are described in Section J in S1 Appendix.

#### Regional differences in whooping cough incidence

Aggregating provincial whooping cough data to the national level (Fig 8) reveals a pattern consistent with previous analyses that lacked provincial data [22]. One feature of this pattern is an apparent resurgence of whooping cough in the 1990s (highlighted in Fig 8). We find that this much-discussed resurgence (e.g., [22]) was not uniformly expressed across the country (Fig 8 bottom six panels), and is clearly apparent only in the territories, the prairies, and Qúebec. Throughout the 1990s, yearly cases per 100, 000 peaked at 21 in Ontario, but in the territories the peak was 293. The original study [22] did not have the sub-national data required to explore these regional differences.

**Fig 8.**
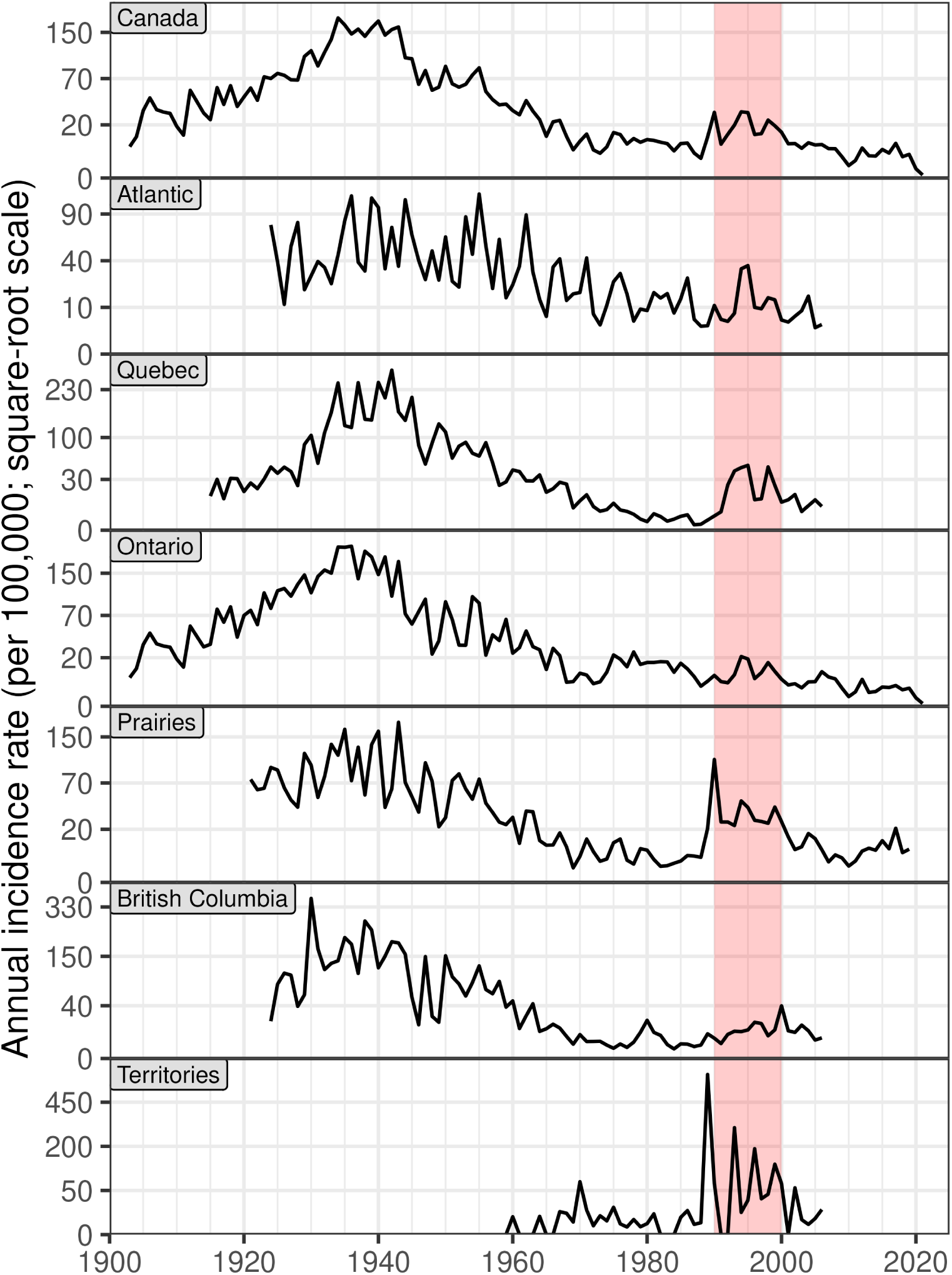
Regional[23] differences in average annual whooping cough incidence in Canada over twelve decades (square root scale). The national data (top panel) are very similar to the first figure in a review based on different data sources [22]. The red region (1990–1999) highlights the first resurgence in national whooping cough incidence since widespread vaccination began in 1943. This plot shows that not all regions peaked in the 1990s, a pattern that could not have been detected with the national data used in the original study. Methods for producing this plot are described in Section K in S1 Appendix.

## Discussion

CANDID complements existing Canadian notification data. The Public Health Agency of Canada (PHAC) provides an online portal [10] (https://diseases.canada.ca/notifiable) with annual, national incidence counts often used in retrospective analyses (e.g., [22, 24–28]). However, these data lack the detail needed to study outbreak patterns, seasonality, or geographic variation. Our sub-annual, sub-national data enable research on patterns of variation within years and across provinces in Canada, and facilitate comparisons with U. S. data [29]. While Public Health Ontario provides online monthly data since 2012 [30], our archive includes weekly Ontario data (1990–2021) and extends back before 1924, including Ontario (1903), Saskatchewan (1910), and Qúebec (1915). By consolidating federal and provincial sources (Section A in S1 Appendix) into a standardized format (Sections D-G in S1 Appendix), we simplify integration of new data, enabling researchers to focus on analysis rather than curation.

Our project parallels Project Tycho [31], which curated weekly U. S. incidence data and whose impact is summarized by [29]. Unlike Tycho, we open-sourced our data preparation pipelines to enable community-driven quality improvements. These pipelines include scans of original documents, spreadsheet replicas, and scripts to convert them into tidy CSV files. To our knowledge, no other studies publish such replicas (see the Data entry section), which help identify and correct data-entry errors. Our open-science approach enables researchers to trace incidence counts back to original sources (Section H in S1 Appendix) and improve data quality over time.

A well-established feature of poliomyelitis is its seasonal pattern, with epidemics in North America typically occurring in summer. Ref. [11] attributed earlier epidemics in southern U. S. states to higher transmission rates. Our data extend this latitudinal gradient northward: epidemics across Canada tended to occur later (August–October) than those in the U. S. (May–July). However, we find no substantial differences in timing among Canadian provinces from 1934 to 1960, despite large variation in climate.

### Future directions

In this initial release of CANDID, we focused on sub-annual and sub-national disease incidence, with plans to expand further. First, we will include data stratified by age, sex, and municipality for available time ranges, provinces, and diseases. Age data, for example, can be critical for estimating the impact of vaccination programmes in childhood diseases (e.g., [32]). Second, we will extend the dataset’s time range, disease coverage, and geographic detail as finer-scale or corrected data become available. Third, we will curate population-level information useful for epidemiological analyses, such as birth rates, mortality, vaccination, and school-term dates. Beyond expansion, we will address open questions in Canadian infectious disease history. For example, we will test whether regional differences in vaccination programmes could explain spatial variation in the size of the 1990s resurgence of whooping cough (Fig 8). Finally, we will explore AI-powered optical character recognition, using our archive as a training dataset to enhance its efficiency and accuracy.

### Logistical challenges

The process of assembling CANDID highlighted logistical obstacles that are likely to arise in other countries seeking to build comparable historical surveillance datasets. The first challenge was locating and obtaining source material. Historical records were dispersed among federal and provincial publications issued under evolving titles and formats, with many available only as hard copies in archives or libraries. Coordinated searches, correspondence with data stewards, and formal data requests to multiple agencies and libraries were required. These challenges are described in more detail in Section A in S1 Appendix.

A second challenge was manual data entry. Optical character recognition could not accurately extract numerical tables from scans, necessitating full manual transcription. Coordinating several data enterers required standardized templates, shared conventions for unclear entries, and frequent communication to ensure consistency across spreadsheets. Looking ahead, advances in artificial intelligence may mitigate some of these difficulties, as noted in our future plans (see the Future directions section), although many questions remain.

A third challenge was harmonizing information drawn from sources that differed widely in format and structure. Converting these diverse formats into consistent, machine-readable CSV files and resolving overlaps across locations, time scales, disease hierarchies, and data sources required careful sequencing of processing steps and version control across multiple repositories. These challenges are detailed in Sections D-G in S1 Appendix.

Finally, quality control involved iterative cross-checks across diseases, jurisdictions, and time scales. Addressing discrepancies meant revisiting earlier stages of data entry and processing, emphasizing the importance of reproducible pipelines. These challenges illustrate the logistical scale of constructing a comprehensive historical surveillance archive. By documenting our workflows and tools openly, we have worked for CANDID to serve as both a resource and a practical starting point for researchers undertaking similar digitization efforts elsewhere.

### Limitations

Under-reporting is a known limitation of surveillance data [31, 33]. Correcting for it requires disease- and context-specific methods that rely on supplementary data (e.g., serological surveys, case-fatality ratios, demography) and modelling [34]. Given the number of diseases and years covered, such corrections are beyond our scope. Our contribution is to make data available in a form that supports future work, including efforts to address under-reporting. As one step in that direction, we distinguish true zero case counts from data that were unreported, based on information available in the original sources (see Section E in S1 Appendix).

In addition to under-reporting, two other factors complicate the interpretation of these incidence data: evolving sub-disease hierarchies and inconsistent time scales. First, changes in how diseases are classified and reported over time affect comparability, as the level of aggregation can vary year to year (e.g., the normalized dataset includes 37 meningitis sub-diseases, with 1–15 reported in any given year; Figs C-F in S1 Appendix). Second, variation in reporting frequency (e.g., weekly, monthly, quarterly; Fig 4, Fig 5, Fig 6) hinders the construction of evenly spaced time series, which are often needed for modelling and visualization.

Historical gaps in CANDID persist due to surveillance program changes (e.g., chickenpox was not notifiable from 1959–1985) and incomplete source coverage, particularly outside Qúebec, Ontario, and Saskatchewan before 1924. These gaps range from missing weeks (e.g., lost book pages) to illegible handwritten records (see the Quality control section). Although transcription and coding errors cannot be ruled out, we compared reported subtotals with their marginal totals and corrected all identified discrepancies for our primary example diseases (whooping cough, poliomyelitis). By releasing open data pipelines, we aim to enable similar checks across all diseases and to support collective efforts to improve the completeness and reliability of CANDID over time (details in Section I in S1 Appendix).

## Conclusion

More than a century of infectious disease surveillance in Canada has produced a valuable record of epidemic patterns that has been largely unexploited, but can now be easily accessed. Comprehensive sub-annual and sub-national Canadian infectious disease incidence data have previously been unavailable. Similar data from other countries have been critical to establishing the foundations of epidemiological modelling and continue to push the field forward (e.g., [1, 35–42]).

CANDID makes it possible to study variation in disease incidence within years and across provinces, with applications ranging from infectious disease research to broader interdisciplinary work. For example, it can be used to assess how incidence in Canada relates to socio-economic factors such as urbanization and wealth inequality. The dataset also supports public health planning by situating recent outbreaks and epidemics within their historical context.

In principle, it should be straightforward to keep the archive up to date, but doing so will require the cooperation of provincial and territorial public health agencies, which have released *less* data publicly since strictly digital data collection began in the 1990s. We contacted all these agencies but were able to obtain recent weekly data from only two provinces. While we recognize that agencies may face practical constraints, such as the staff time required to prepare and maintain public releases, we encourage Canadian governments to support routine public access to weekly, aggregated counts of infectious disease notifications.

## Data Availability

All data produced are available online at https://github.com/canmod/iidda

## Supporting information

### S1 Appendix Methodological details

This supporting information details the historical and methodological framework for compiling and preparing Canadian notifiable infectious disease incidence data. It explains the sources and evolution of the Canadian Notifiable Disease Surveillance System (CNDSS), including federal and provincial publications, and describes the process of locating, digitizing, and cleaning these historical records. Data entry procedures are described along with the individuals who did the work. Quality control procedures are described, including internal consistency checks across time scales, locations, and disease hierarchies, as well as comparisons among data sources. Finally, the document outlines specific analytical methods for diseases like polio and whooping cough.

## Acknowledgments

We are deeply thankful to Alberta Health and Public Health Ontario for providing us with recent weekly incidence data. Research assistants Jen Freeman, Frank Jin, Ronald Jin, and Steven Lee wrote code that we used during this project. Research assistants Jeanne Lin, Saul Widrich, Qinxian Zhu, Claire Lees, and Julia Maja entered some of the data. Research assistants Maya Earn, Arielle Earn, and Elizabeth O’Meara found, organized, and scanned source documents. We appreciate the enthusiastic encouragement we received from Caroline Colijn, Michael Li and many other colleagues in the Canadian Network for Modelling Infectious Diseases (CANMOD).

## S1 Appendix: Methodological details

### Supporting Information for

#### A Data Sources

In 1924, the Dominion Bureau of Statistics, now Statistics Canada, began collecting communicable disease incidence data.

The main objectives of this system are to provide a mechanism for monitoring the health of the population by identifying and responding to changes in reporting trends of specific diseases and to provide information that can contribute to the development of health policy and the planning of care, prevention, and control programs. [1]

In 1988 this initiative was transferred to the Canadian Laboratory Centre for Disease Control and persists today under the administration of the Public Health Agency of Canada (PHAC), as the Canadian Notifiable Disease Surveillance System (CNDSS) [2]. Data for the CNDSS are provided by provincial and territorial governments to monitor diseases of public health concern [2]. Prior to the onset of this federal initiative in 1924, several provinces were already collecting such data without reporting it to the federal government.

Our group’s search for historical Canadian infectious disease notification data began in 2000. Initially, D. J. D. Earn acquired handwritten weekly counts for Ontario, covering five decades (1939–1989), from the Ontario Ministry of Health (main text Table 1). A significant breakthrough occurred during a visit to the chief medical officer of health in Manitoba, where he discovered a single page of notifications submitted to the Dominion Bureau of Statistics. He photocopied this document and sent it to Statistics Canada, as it was the first evidence that the data tables we were seeking existed.

In 2002 and later, D. J. D. Earn engaged in extensive communications with Statistics Canada via telephone and email. As a result, they provided photocopies of handwritten weekly and monthly notification spreadsheets from 1924–1955 that they had located in their archives. At the time, our resources were insufficient to digitize and clean all of these data. However, we did present analyses of a few disease time series in publications (e.g., [3, 4]). Additionally, we located more published CNDSS data from 1956–2000 in University Libraries and from Statistics Canada. During this historical period, the publication of data transitioned from weekly to monthly and eventually to quarterly. In 2021, with funding from the Canadian Network for Modelling Infectious Diseases (CANMOD), D. J. D. Earn initiated a systematic effort to complete this 2+ decade project to locate, digitize, clean, and distribute all available CNDSS data.

The following paragraphs describe the sources of data, including both federal and provincial government compilations. Throughout the project, D. J. D. Earn, G. MacKinnon, S. Manzin, C. Shi, A. Earn, E. O’Meara, and S. C. Walker contributed to identifying and locating these sources.

##### Federal government compilations

The CNDSS data have been published using different titles that have changed over time, many of which are (at the time of writing) difficult or impossible to find using Google without physically going to libraries and archives. Ref. [5] provides annual communicable disease incidence data by province from 1924 to 1954, but it does not provide a list of the titles used to publish these data. We have been able to find some of these titles, but not all of them. The titles used by Statistics Canada and Health and Welfare Canada to publish CNDSS data changed over time, and we have been able to find some of these titles, but not all of them.

**1924-1959** During this period, the Dominion Bureau of Statistics compiled provincial communicable disease incidence data. Photocopies that we obtained from Statistics Canada are evidence of these compilations. In 1954, the Bureau published annual incidence data from 1924-1952 [5]. This report states that the Dominion Bureau of Statistics began publishing incidence data in 1952 under the title “Summary of Cases of Notifiable Diseases in Canada”. This report also states that the Dominion Bureau of Statistics compiled “a restricted monthly release” of incidence data from 1924–1932 and that in 1933 this became a “printed weekly report.” The phrase “weekly communicable disease report” was used by Ref. [5], suggesting that this was the name of the “printed weekly report”. Again we were unable to find these releases, reports, or publications on the internet, but we have photocopies of data that were presumably used to produce them.
**1959–1974** From 1959-01-24, the Dominion Bureau of Statistics published weekly provincial CNDSS data under the title “Notifiable Diseases – Weekly Summary” until they were renamed Statistics Canada on May 1st 1971. Statistics Canada continued using this name until 1974. Many of these reports from 1959 to 1974 can be found in the Research & Collections Resource Facility at the University of Alberta.
**1975–1976** Statistics Canada and Health and Welfare Canada jointly published weekly provincial CNDSS data using the title “Notifiable Diseases Weekly Summary Provi-sional Report” for less than one year from 1975-04-26 to 1976-01-03.
**1976–1978** During this period, Health and Welfare Canada published weekly provincial CNDSS data using the previous title “Notifiable Diseases – Weekly Summary”.
**1979–1989** During this period, Statistics Canada and Health and Welfare Canada jointly published 4-weekly provincial CNDSS data using the title “Notifiable Diseases Summary”. At the time of writing, we were not able to use Google to find a digitized copy of CNDSS data for the first quarter of 1979. However, the Canada Diseases Weekly Reports (CDWR) reproduced CNDSS data using this title starting with the four-weekly period ending 1979-04-21. The CDWR continued to reproduce CNDSS data using this title until 1991, but they did not do so for every four-weekly period. We were able to obtain data for more of these four-weekly periods directly from Statistics Canada, but we were not able to obtain all of them.
**1990–2006** From 1990–1992, Health and Welfare Canada published monthly provincial data under the title “Notifiable Diseases Summary” and continued to publish quarterly provincial data under the same title until the last quarter of 2003 at which time the Public Health Agency of Canada took over until 2006. Many of these publications are reproduced in the Canada Communicable Disease Report (CCDR).
**2007–present** Unfortunately, we were unable to find any sub-annual and sub-national data source covering all provinces after 2007. Since 2001, the Public Health Agency of Canada (PHAC) has maintained a web portal [2] that provides annual, national incidence.

##### Provincial government compilations

Provincial public health agencies have published data directly, and many of these publications predate the federal programme that began in 1924. These sources also varied in the titles under which they published. The following is a non-exhaustive list of titles that have been used by provincial health agencies to publish monthly disease incidence data for their province.

**Ontario, 1903–1905** “The Sanitary Journal of the Provincial Board of Health of Ontario”.
**Ontario, 1906–1924** “Annual Report of the Provincial Board of Health”.
**Ontario, 1925–1947** “Annual Report of the Department of Health”.
**Saskatchewan, 1910** “Annual report of the Bureau of Public Health for the Province of Saskatchewan”.
**Saskatchewan, 1921–1922** “Annual report of the Bureau of Public Health of the Province of Saskatchewan”.
**Saskatchewan, 1923–1926** “Annual report of the Department of Public Health of the Province of Saskatchewan”.
**Saskatchewan, 1927** “Annual report of the Department of Public Health and the Vital Statistics report of the Province of Saskatchewan”.
**Québec, 1915-1922** “Rapport annuel du Conseil supérieur d’hygiène de la province de Québec = Annual report of the Superior Board of Health of the Province of Quebec”.
**Québec, 1923-1934** “Rapport annuel du Service provincial d’hygiène de la province de Québec = Annual report of the Provincial Bureau of Health of the Province of Quebec”.
**Manitoba, 2004-2017** “Manitoba Monthly Surveillance Unit Report” – available online (at the time of writing) here: https://www.gov.mb.ca/health/publichealth/surv eillance/episummary/archive.html.

##### Provincial and territorial government data requests

In 2021, we began reaching out to provincial and territorial public health agencies for more recent sub-annual data. Although we corresponded via email with all such agencies, only Public Health Ontario and Alberta Health provided us with recent data. Both agencies provided us with weekly data. This work was carried out by G. MacKinnon and S. Manzin, under the supervision of S. C. Walker.

#### B Data entry

Data were manually transcribed from scanned reports into standardized Excel spreadsheets following project digitization guidelines:

https://github.com/canmod/candid/blob/main/digitization_process.md

The core principle of this document is that digitized spreadsheets should mirror the structure and appearance of the original source documents closely enough for easy comparison, while allowing flexibility when an exact match is impractical. In practice, maintaining consistency across data enterers required more than simply following this principle. Perfect replication was sometimes unrealistic because capturing every small detail demanded more time and effort than available, requiring decisions about which details were worth the effort. In other cases, readability was intentionally improved through small departures from the source, such as alternating row shading. At times, Excel itself could not reproduce the layout of the original–for example, when a number appeared between two table cells–necessitating collective decisions about how to represent such cases in a spreadsheet. The guidelines therefore serve as a shared reference to help data enterers make similar judgments in these situations.

Regular meetings among research assistants were used to resolve ambiguities and update the document. The main team of data enterers included S. Manzin, G. MacKinnon, C. Shi, J. Lin, Q. Zhu, C. Corradi, S. Widrich, J. Maja, and C. Lees. Manzin, MacKinnon, and Shi entered the largest volume of data, with Lees contributing substantial work prior to the start of the systematic digitization effort in 2021. Their work included manual entry from scanned tables into spreadsheets, checking and correcting data entry errors, and scanning hard-copy source documents. Each spreadsheet contains the name of the data enterer in a dedicated cell.

The guideline document is organized around reusable Excel templates that promote consistency across similar sources and formats; these templates contained table structures with row and column headers but no data values, allowing them to serve as standardized starting points for digitizing sources with recurring formats. Some sources were digitized quickly by a single assistant, making separate documentation unnecessary because the resulting spreadsheets themselves defined the format; as a result, not all data sources are represented in the document.

#### C External links

Download links to the datasets described in this paper, as well as others from our broader Canadian historical epidemiological data digitization project, can be found at the following web page:

https://github.com/canmod/iidda/blob/main/README.md

In addition to download links, which page covers the following additional technical information:

- **Data Dictionary**
- **Data Harmonization**
- **Reproducing IIDDA Datasets**, including:

**–** Running natively
**–** Running in a Docker container
**–** Running interactively
**–** Dependency management
**–** Requirements
- **Project Structure**, including:

**–** Data sources and pipelines (source data and source code)
**–** Derived data and tidy datasets
**–** Identifiers
**–** Metadata
**–** Lookup tables
- **Contributions**, including:

**–** Contributing source data and pipelines
**–** Contributing fixes to data and pipelines
**–** Contributing to IIDDA project development

We have also written a small package for reading the data directly into R. An introduction to this package can be found here:

https://canmod.github.io/iidda-tools/iidda.api/articles/Quickstart

This iidda.api package is part of a suite of packages including tools that are used in data preparation pipelines. The source code for this suite is available here:

https://github.com/canmod/iidda-tools

All the code to produce the figures and statistics in this paper are available at:

https://github.com/canmod/candid

#### D Data preparation pipelines

Each data preparation pipeline followed one of the paths outlined in Fig A. Data sources were provided as hard copies, digitally produced PDF files, or digital spreadsheets. Digital spreadsheets were particularly advantageous because they enabled us to directly script the production of CSV files. When data were not in spreadsheet format, conversion was necessary before scripting could begin. For digitally produced PDF files, we tried to use automated tools like PDFTables (https://pdftables.com/) to convert them into spreadsheets. Hard copies, however, always required scanning followed by manual data-entry into spreadsheets. We were unable to find a viable optical character recognition (OCR) approach to avoid manual data-entry. However, in the years since we began this systematic effort to digitize Canadian incidence data there have been tremendous advances in artificial intelligence (AI), and so this situation may have changed. In future work on digitization we plan to replicate parts of this work using OCR, to test it as an efficiency tool in this area.

All CANDID preparation pipelines are available at:

https://github.com/canmod/iidda/tree/main/pipelines

The main scripts for producing each of the three datasets (unharmonized, harmonized, and normalized) discussed in this article are available at:

https://github.com/canmod/iidda/tree/main/pipelines/canmod-compilations/prep-scripts

These scripts depend on outputs produced by other pipelines that generate data from specific sources. Instructions on how to reproduce all of these outputs are provided in the README.md file on this GitHub repository:

https://github.com/canmod/iidda

The data preparation pipelines were written by G. MacKinnon, S. Manzin, F. Jin, and S. C. Walker. These individuals, in addition to S. Lee, R. Jin, M. Roswell, and J. Freeman, also wrote software packages that were used in these pipelines. The reason for separating pipelines from packages is that pipelines are specific to this project, whereas packages are designed to be reusable in other digitization projects. The packages used in these pipelines are available at:

https://github.com/canmod/iidda-tools

#### E Preparing unharmonized CSV files

We developed one open-source R script for each spreadsheet that converts it into a tidy CSV file [6]. These R scripts used the unpivotr package [7] to convert the wide-format data used in historical documents to long-format data that make it easier to manipulate using standard tools [6]. Long-format data also make it easier to combine data from different sources into a single data set, by ensuring they each consist of fields from the same standardized data dictionary. All of the resulting CSV files have been collected into a single file that we label *unharmonized* because it contains unharmonized historical disease and place names.

**Fig A:**
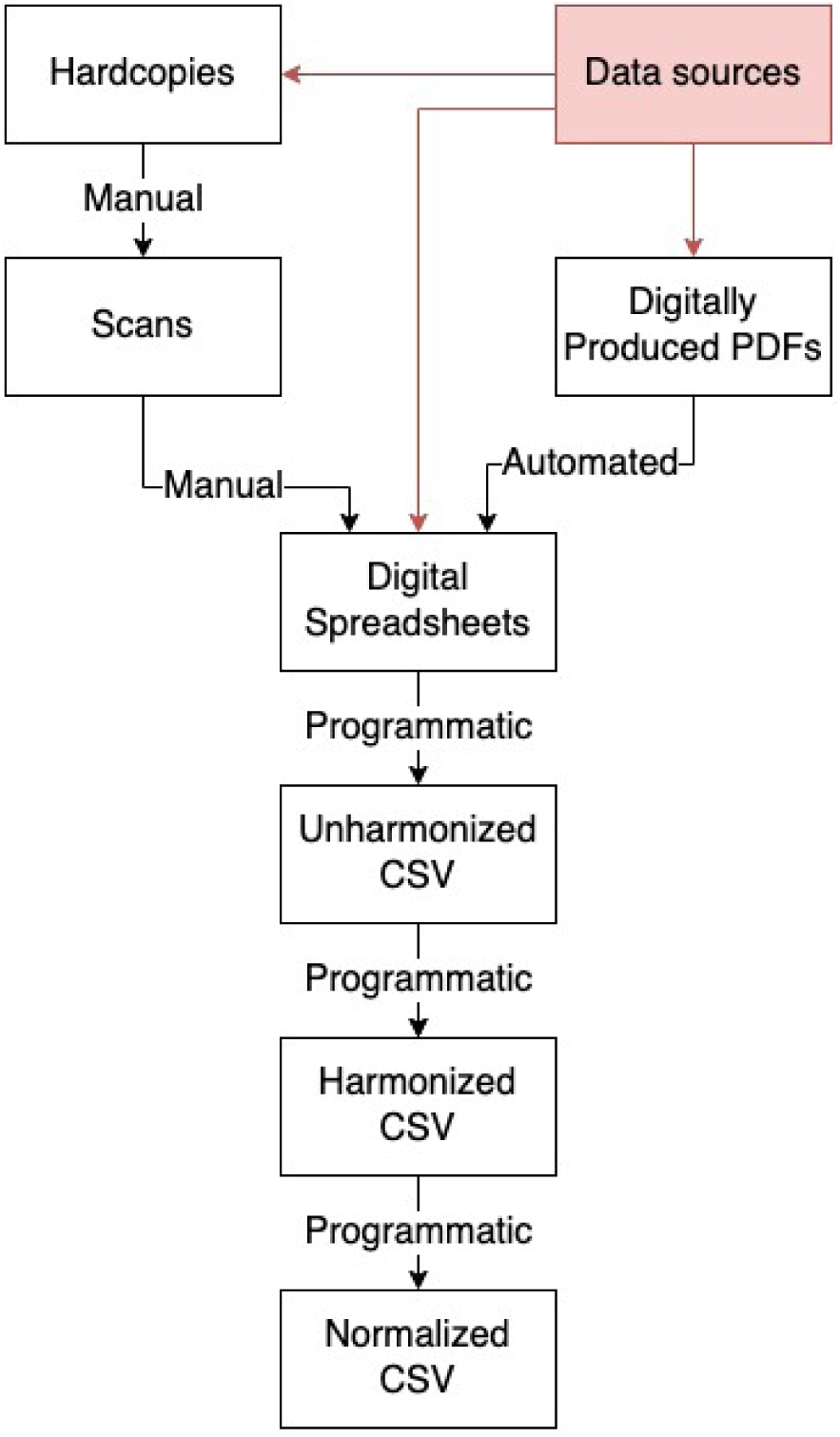
Data preparation pipeline overview. All products begin with data sources (red), which provide hard copies, digital spreadsheets, or digitally produced PDFs. Processing steps are classified as manual (e.g., data-entry), automated (e.g., PDF table extraction tools), or programmatic (e.g., customized R script).

These unharmonized data also include information on why certain incidence values are not available, when we could determine the reason from the sources. In the column containing numbers of cases, cases_this_period, we allowed the following types of values:

- Non-missing numeric case numbers (non-negative integers).
- One of the following strings explaining why the case numbers are missing^1^:

**–** The phrase ‘Not available’, for unknown reasons.
**–** The phrase ‘Not reported’, for unknown reasons.
**–** The phrase ‘Not reportable’, presumably indicating that the jurisdiction was not required to report these numbers.
**–** The word ‘Missing’, typically indicating missing pages in the middle of a multi-page table.
**–** The word ‘Unclear’, meaning that the value is missing from CANDID because it is not legible.
**–** The word ‘Unclear’, with a special string format,

`{guess_1} - {guess_2} - … - {guess_n} (unclear)`

(e.g., ‘`36-23-59 (unclear)`’), meaning that the value is missing because the number is difficult to read but we have one or more guesses. In the harmonized datasets we use the first (i.e., best) guess (see the section on Preparing harmonized CSV files).

**–** Phrases of the format ‘`Wrong but clear total in this cell is {value}`’, mean-ing that this cell should contain a marginal total (e.g., annual total), and that the value is clearly written, but is not the correct total obtained by summing up the component values (e.g., weekly counts).

This list of values for recording missing and unclear data is also described in our living document of data entry processes, introduced in the Data entry section.

#### F Preparing harmonized CSV files

We developed one open-source R script for each data source that joins the unharmonized data with harmonized disease and place names. This was achieved by creating lookup tables containing all historical names, and then adding columns for harmonized names. Links to these lookup tables are available at:

https://github.com/canmod/iidda/blob/main/README.md#canmod-digitization-project

Because sources sometimes reported diseases hierarchically, our harmonized disease names were provided in two columns: `disease`, giving the name of the disease being reported, and `nesting_disease`, optionally giving the name of another disease within which the `disease` is nested. We refer to the values in these `disease` and `nesting_disease` columns collectively as ***disease names***. These disease names are organized into hierarchies, such that most disease names are nested within another disease name (e.g., hepatitis-A is nested within hepatitis-A-B). We refer to disease names that are not nested within any other disease name as ***basal diseases***. These basal diseases are plotted in Figures 4-6 of the main text. For a given combination of location and time period, all disease names at or below a specific basal disease in the hierarchy are referred to as its ***sub-diseases***.

In addition to joining harmonized names, the harmonization scripts also apply the following changes:

- Apply fixes to dates and locations that were obviously entered incorrectly in the original source documents (e.g., one whooping cough record from 1943 was for a week ending on a Sunday, while all other data from the same source were for weeks ending on a Saturday).
- Aggregate data that were stratified by age or city.
- Replace alternative characters for reporting zero cases with a literal 0 (e.g., often a dash was used).
- Remove records containing missing values or text strings indicating the type of missing value (e.g., ‘Unclear’, ‘Missing’).

These fixes add convenience at the expense of removing information contained in the original source, but this information remains accessible in the unharmonized data.

We collected all of the resulting harmonized CSV files into a single file that we label *harmonized*.

#### G Preparing the normalized CSV file

We developed an open-source R script to remove overlapping data and to add data that are implied but not explicitly provided by the original source documents. Our goal was to ensure that each apparently reported case is represented by a single incidence value in the resulting CSV file. We refer to the resulting file as *normalized* aligning with the concept that normalized databases represent each ‘fact’ only once [6].

There are five sources of overlap that could cause cases to be counted more than once:

- Locations (e.g., provincial data being reported along with national data)
- Data sources (e.g., weekly data for Ontario between 1939–1978 being reported by both Statistics Canada and the Ontario Ministry of Health)
- Time periods (e.g., weekly data being reported along with monthly data)
- Disease hierarchies (e.g., polio with and without paralysis being reported along with total polio)
- Mixtures of the previous sources of overlap (e.g., some provinces have only monthly data, while others have only weekly data.)

In addition to removing overlapping historical records, we also add records that are implied by the information in the sources. There are two types of implied information in the harmonized data that we have made explicit in the normalized data:

- Unaccounted cases that are detected when the reported number of cases for a disease is greater than the total over its sub-diseases.
- Missing data at finer time-scales (e.g., weekly, monthly) can be assumed to be zero if zeros are reported at a coarser time-scale (e.g., yearly data) that temporally bound the finer scale.

The CANDID archive curates data from a wide variety of sources (main text Table 1) and diseases, each with different biases and quality issues. It is therefore impossible to produce a perfectly normalized dataset, nor is it our goal to do so here. Instead we aim (1) to make reasonable choices so that the analyses in this paper respect basic principles of consistency (e.g., each case is counted at most once), (2) to set up a data processing pipeline that allows sustained work to improve the quality of the normalization process, and (3) to make the complete and un-normalized data easy to access so that others with expertise in a particular area can make improved normalization choices.

In the remainder of this section, we describe the normalization steps that we take to produce our posted normalized files. Users are free to modify these steps by modifying the scripts and other files these scripts depend on (see the Data provenance section for information on how to find these scripts and files).

##### Add unaccounted cases

Sometimes the total for a `nesting_disease` is reported along with some, but not all, of its sub-diseases. In these instances, after having ruled out other known data quality issues, we produced records with incidence counts given by the associated reported total minus the sum of the reported sub-diseases. These incidence values can be identified in the normalized dataset by a value in the `disease` column of the form `nesting-disease_unaccounted`, and with `derived-unaccounted` in the `record_origin` column. These sums were computed by grouping by time-period, province, data source ID, and `nesting_disease`.

##### Join population data

The estimated provincial population for each incidence value was joined to the normalized dataset. As a result, population numbers are repeated in the dataset because incidence values for different diseases are linked to the same population size within a specific period and province. While this repetition technically violates our normalization principle, the added convenience justifies this step. The population column provides linearly interpolated estimates of the intercensal populations for each province at the mid-point of each period, using census-derived data from [8–10].

##### Resolving overlapping locations

Filtering out all data for the entire country easily solves this source of overlap.

##### Compute implied zeros

In some instances, there are zeros reported at a coarse time-scale (i.e., for a year), but the data at a finer timescale (weekly/monthly) for the same disease and location is empty or not available. We replaced weekly data that were missing and/or not available in national data sources with zeros when they were implied by a zero at a coarser timescale for the same disease and location. These incidence values can be identified in the normalized dataset by a value of derived-implied-zeros in the record_origin column. These implied zeros were given lower priority than other weekly data when resolving overlap, as we describe next.

##### Resolving overlapping data sources and time-scales

We generally prioritize national data sources that report for all provinces (e.g., Statistics Canada) over provincial data sources that report for a single province (e.g., Saskatchewan Bureau of Public Health). We always prioritize finer time-scales (e.g., weekly) over coarser ones (e.g., quarterly). For example, if monthly data from a national source overlaps with weekly data from a provincial source, we will choose the weekly provincial data.

We handle data source and time-scale overlap sequentially, starting with an empty dataset and adding records from a dataset produced by applying the previous normalization processes to the harmonized data. At each step in this sequence, we consider new candidate records and only add those that do not overlap temporally with the existing ones. Being added first therefore indicates a higher priority:

1. All weekly data from national sources.
2. Non-overlapping weekly data from provincial sources.
3. Non-overlapping two-weekly data from national sources.
4. Non-overlapping two-weekly data from provincial sources.
5. Non-overlapping weekly implied zeros from national sources.
6. Non-overlapping monthly data from national sources.
7. Non-overlapping monthly data from provincial sources.
8. Non-overlapping quarterly data from national sources.

For any time period and province, we have at most two data sources: one from a federal organization (e.g., Statistics Canada) and one from a provincial organization (e.g., Saskatchewan Bureau of Public Health). To address such overlap when it occurs, we prefer national sources to those from provincial sources. This choice has the advantage of being easy to apply and also has a better chance of producing provincial data streams that are comparable because we can inherit the choices that the federal organization made when publishing data from different provinces.

If the two sources produced identical results then this choice would be irrelevant. Although there are periods for which national and provincial sources reported identical counts, this is not typically the case. Fig B gives an example comparing 37 years of weekly whooping cough data in Ontario as reported by Statistics Canada and the Ontario Ministry of Health. This figure shows that until 1970 the two agencies were reporting virtually identical numbers, with the occasional deviation. In contrast, there are deviations consistently from 1970 to 1977, although the qualitative pattern is still similar.

##### Resolve overlap caused by disease hierarchies

To address how this type of overlap is addressed, it is necessary to further define terminology related to disease hierarchies, building on the concepts introduced in the section on Preparing harmonized CSV files. The ***global hierarchy*** of a basal disease includes all sub-diseases that appear at least once in the harmonized dataset, while the ***local hierarchy*** is specific to a particular location and time period. Some of these global hierarchies are simple (e.g., whooping cough has no sub-diseases at all) whereas others are complex (e.g., meningitis has 37 sub-diseases in the global hierarchy) with local hierarchies changing over time.

We will dig into the meningitis hierarchy a little to give a sense of the complexity. The harmonized dataset contains 33 different local hierarchies of meningitis. The website for the paper has one figure for each of these local hierarchies at this URL: https://github.com/canmod/candid/tree/main/output/disease-hierarchies. Here we plot and discuss four of them (Figs C to F). Each of these figures give the meningitis global hierarchy, highlighting in blue the sub-diseases for a particular local-hierarchy (with all other diseases in red). Disease names starting with “ex” indicate counts that exclude certain disease types. Full disease names are constructed by concatenating node names along the hierarchy with dashes (e.g., meningitis-bacterial-haemophilus-influenza), although some names are abbreviated in the figures to save space. Here we summarize the four local hierarchies illustrated in these figures:

- From 1921 to 1967 Statistics Canada reported a total meningitis count without any sub-diseases (Fig C). The sources give no indication of what kind of meningitis is being reported, possibly because it was not known.
- From 1969–1978 only viral meningitis was reported and this was stratified by coxsackie, echo, and virus-unspecified (Fig D). Even if totals for meningitis-viral or meningitis were given in these sources, they were excluded from the normalized data to avoid overlap.
- From 1979 to 1985 the collection of sub-diseases changed completely to report only meningitis associated with encephalitis (both viral and bacterial, Fig E).
- From 2004 to 2007, Manitoba Health reported a complex collection of sub-diseases (e.g., Fig F). Even in this complex case, there is no overlap in observed diseases in the normalized data.

**Fig B:**
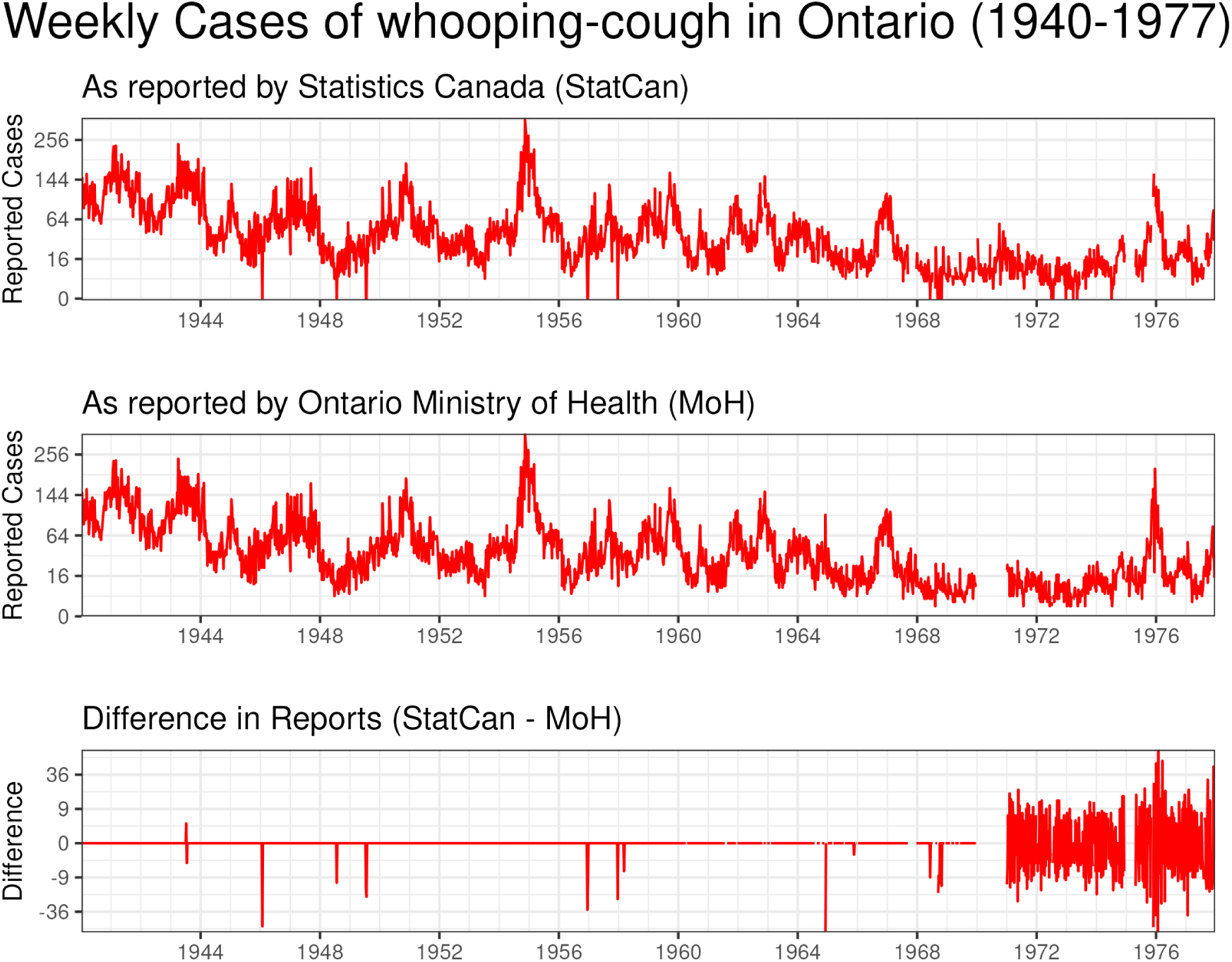
Comparing reported Ontario whooping cough incidence from Statistics Canada with the Ontario Ministry of Health. The difference between the two sources is given on the bottom panel. Additional comparisons among similar data from different agencies can be found on the website for the paper: https://github.com/canmod/candid/tree/main/out put/agency-comparisons

These examples show how we prevent overlap in disease hierarchies. For each basal disease, we keep only the most detailed sub-diseases that were actually reported for a given place and time. Any intermediate or higher-level totals are removed. In effect, all retained sub-diseases are treated as children of the basal disease, ensuring that every case is counted exactly once. For example, Statistics Canada’s 1969–1978 meningitis data (Fig D) included counts for coxsackie, echo, and virus-unspecified meningitis, but also listed a total for viral meningitis. We discarded this total during normalization, so the viral node appears in red to indicate that it is not observed in the normalized data.

For example, the stratification of meningitis illustrated in Fig D came from a data source (Statistics Canada) that also reported a total for viral meningitis (historically called aseptic meningitis), but this total was removed in the normalization process and so the viral node is coloured red to indicate that this sub-disease cannot be observed in the normalized data. As another example, Public Health Ontario reported acute and chronic hepatitis B (Fig G, top), while Statistics Canada and some provinces reported only a single hepatitis B total (Fig G, bottom). Our approach keeps whichever sub-diseases were actually reported locally and removes overlapping intermediate totals.

**Fig C:**
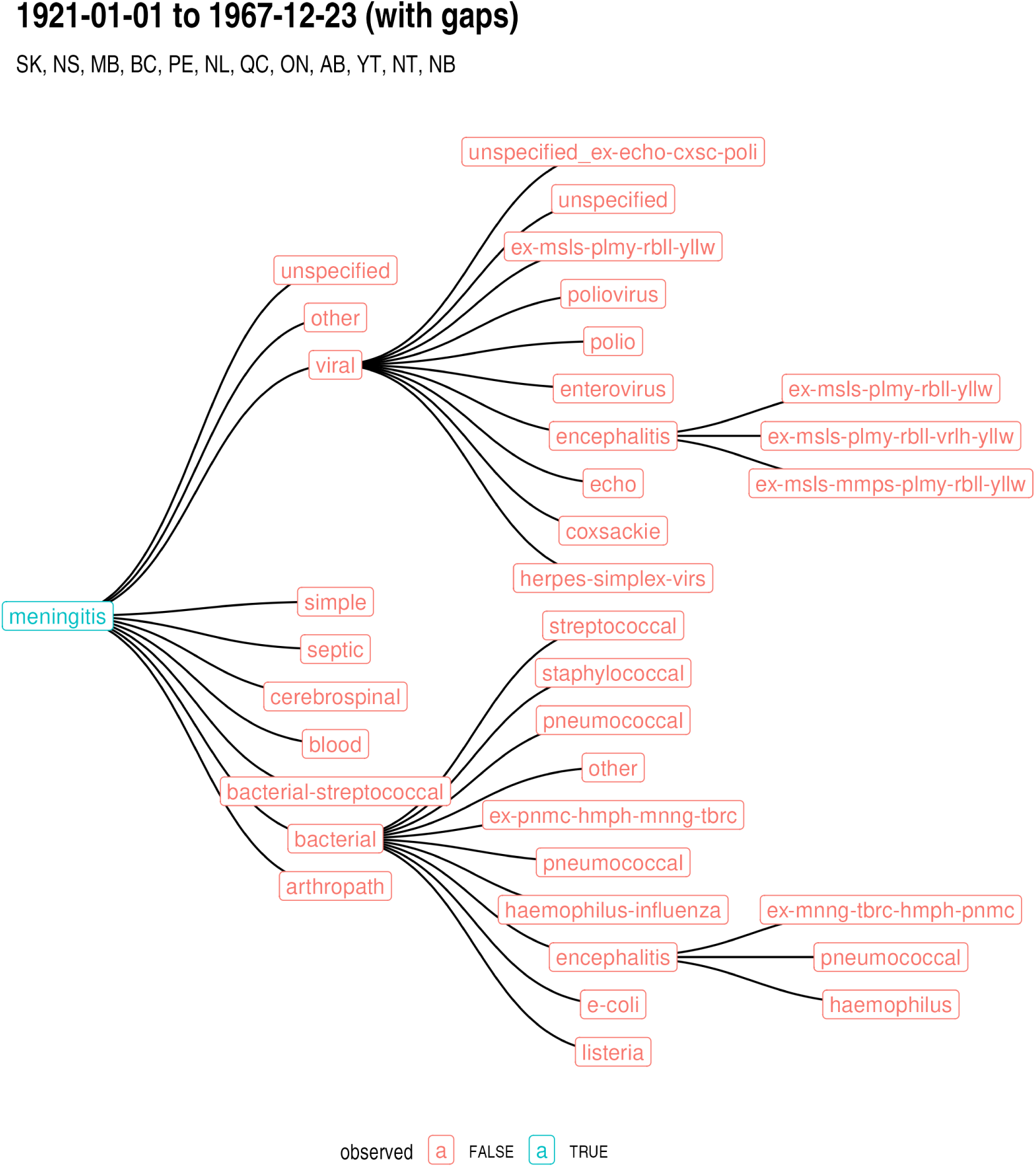
Global meningitis disease hierarchy highlighting in blue a particular local hierarchy of reported sub-diseases between 1921 and 1967. In this local hierarchy, the only reported sub-disease is the basal disease itself.

**Fig D:**
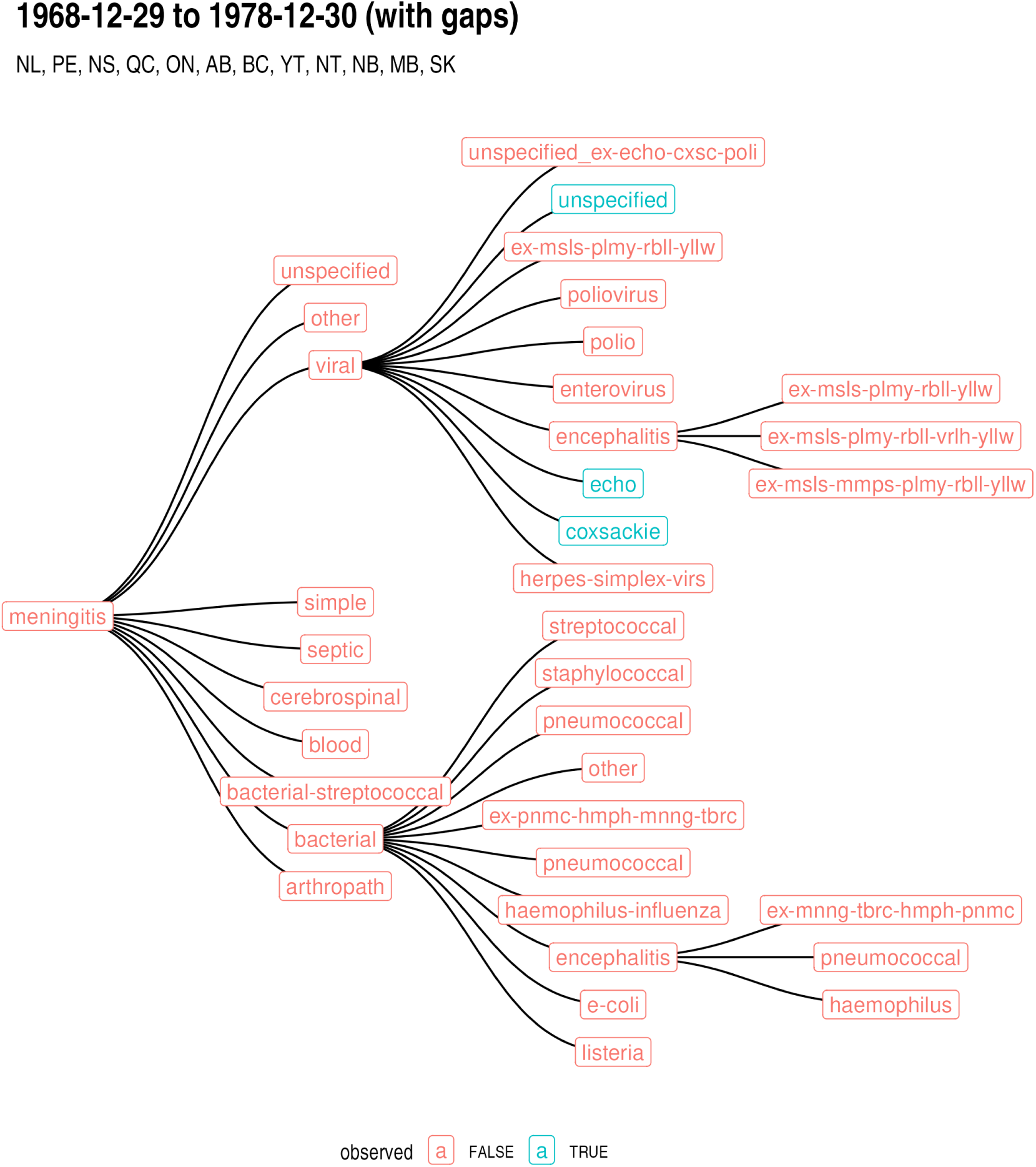
Global meningitis disease hierarchy highlighting in blue a particular local hierarchy of reported sub-diseases between 1968 and 1978.

**Fig E:**
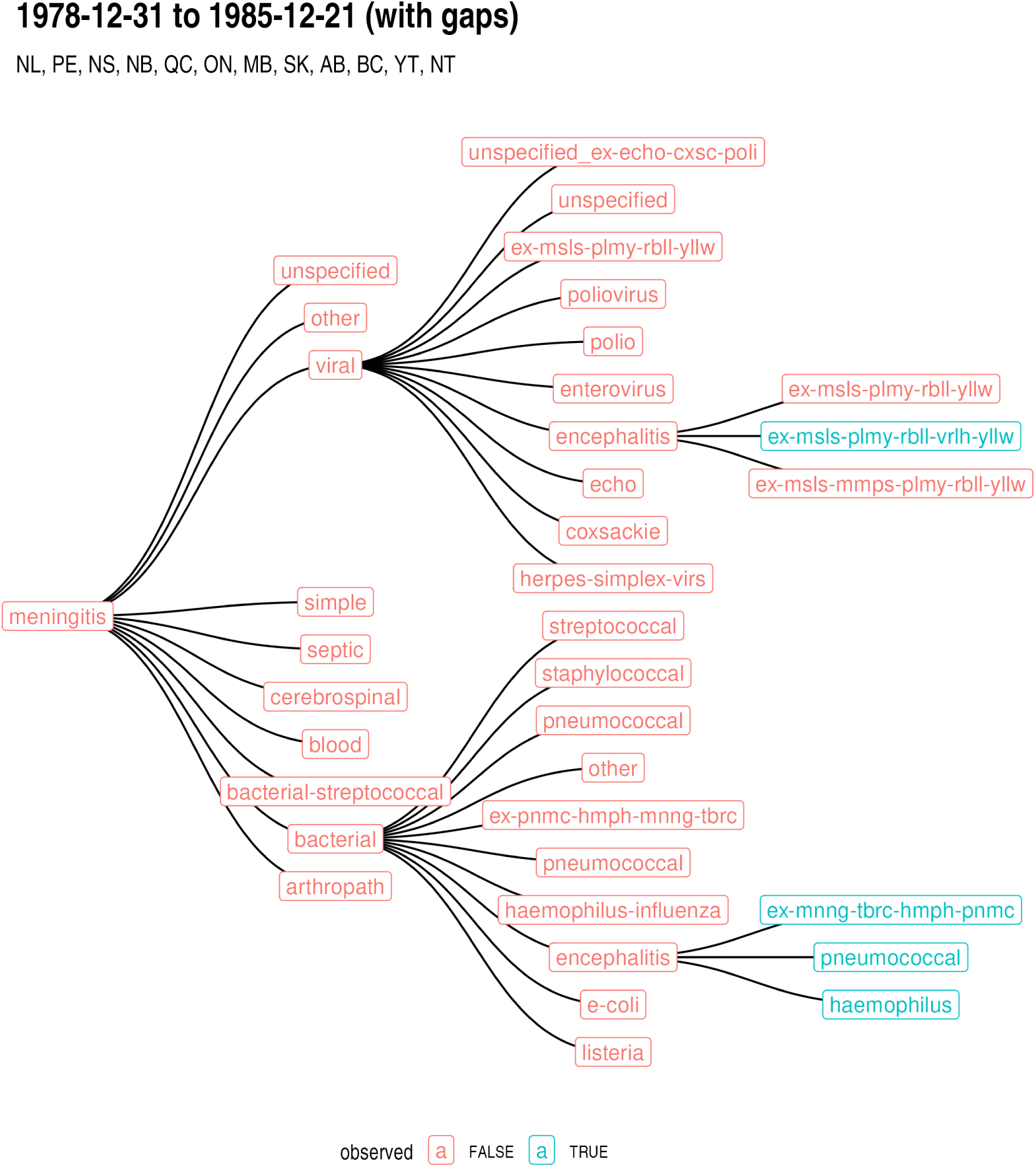
Global meningitis disease hierarchy highlighting in blue a particular local hierarchy of reported sub-diseases between 1979 and 1985.

**Fig F:**
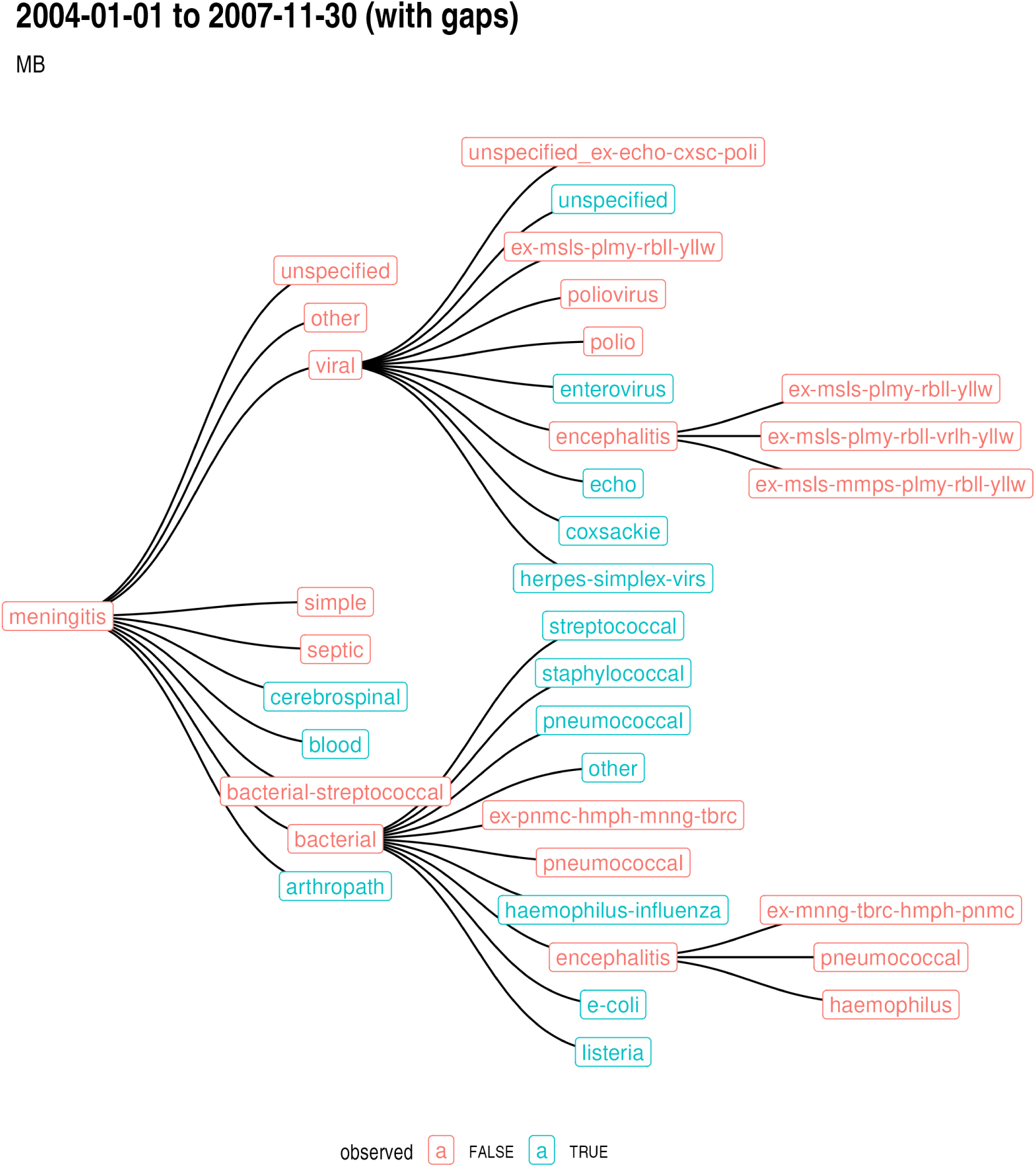
Global meningitis disease hierarchy highlighting in blue the sub-diseases reported by Manitoba Health in the date ranges given.

**Fig G:**
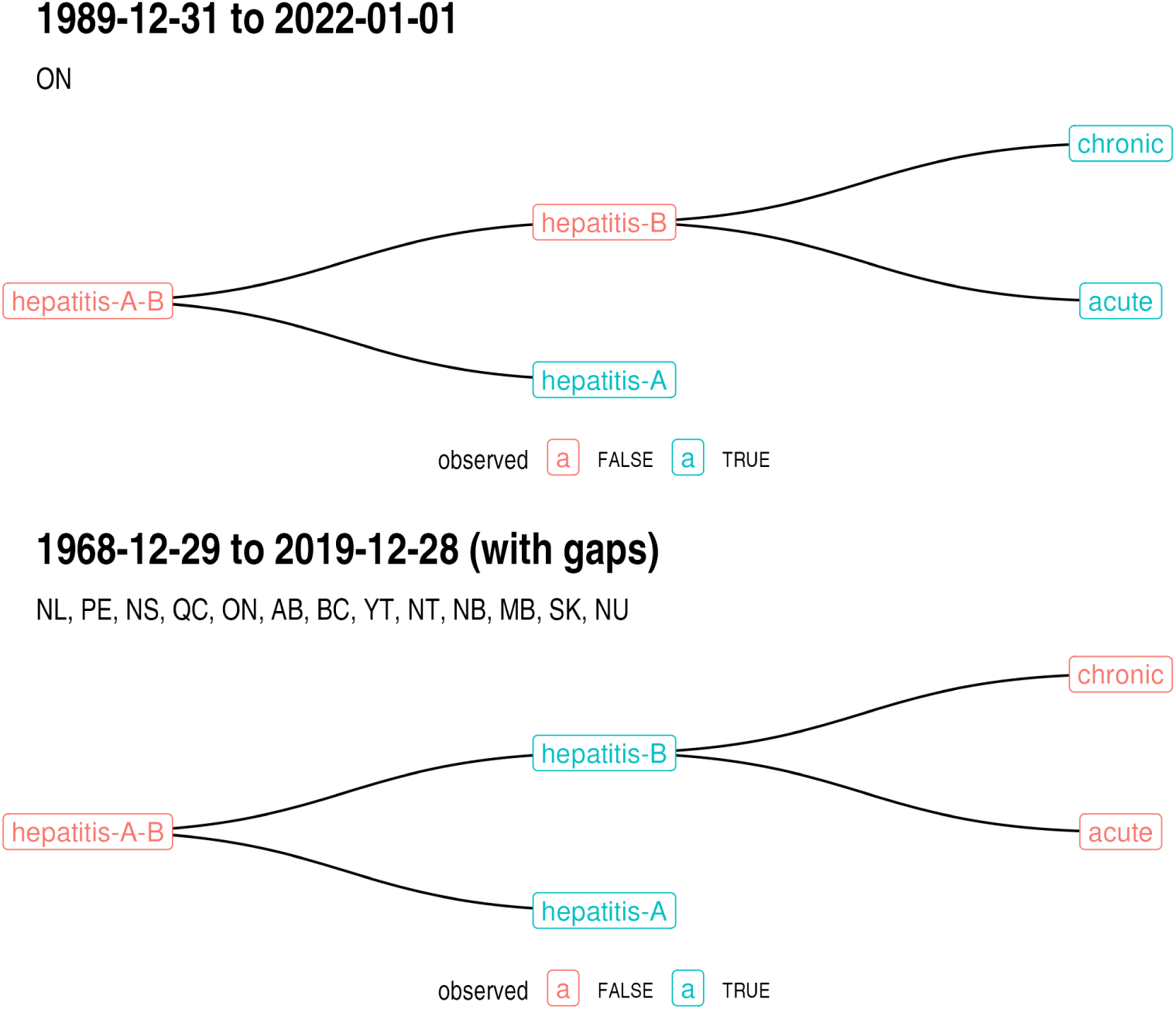
Global hepatitis A and B disease hierarchy, with two panels highlighting different local hierarchies in blue. In the top panel hepatitis B is stratified by acute and chronic sub-diseases, but in the bottom panel hepatitis B is not stratified. See Figs C to F for interpretation details. All hierarchies for hepatitis can be found on the website for the paper: https://github.com/canmod/candid/tree/main/output/disease-hierarchies

##### H Data provenance

The harmonized and normalized data both contain the following columns with unique identifiers to resources used to produce each record:

- `original_dataset_id:` Uniquely identifies the unharmonized dataset containing each record.
- `digitization_id:` Uniquely identifies the digitization (typically an Excel file) containing the data for each record.
- `scan_id:` Uniquely identifies the scan of the original document containing each record.

Records that were implied from associated data (see the section on Preparing the normalized CSV file), as opposed to explicitly reported, do not have entries in the first two of these columns. Data that we received in digital form typically do not have a scan associated with them, and so such records do not have a scan_id.

Information on the above identifiers can be found at:

https://github.com/canmod/iidda/blob/main/README.md#identifiers

The following vignette describes how to investigate the provenance of a record using these identifiers:

https://canmod.github.io/iidda-tools/iidda.api/articles/Provenance

Metadata for the unharmonized, harmonized, and normalized versions of our prepared data are available at:

https://github.com/canmod/iidda/blob/main/README.md#canmod-digitiz ation-project

These metadata contain links to all of the resources (R scripts, Excel/CSV files, and PDFs) used to produce the datasets.

#### I Quality control

We performed two types of quality control checks: marginal total cross checks and comparisons with the PHAC portal. This work was performed by S. C. Walker, G. MacKinnon, S. Manzin, S. Cygu, and Q. Zhu. After describing these two types of checks, we describe how the results of these checks were used to improve the quality of the data by fixing data-entry errors and improving our normalization process.

##### Marginal total cross checks

Using marginal totals from the comprehensive dataset, we performed cross checks over the three available stratifications of data: time scales, locations, and diseases. The analysis involved several comparisons at different levels of data aggregation.

For time-scales, sub-annual records, such as weekly, monthly, or quarterly data, were aggregated to the annual scale and compared to the available annual records. When multiple sub-annual time scales were present, their annual sums were also compared to one another. For locations, sub-national data were aggregated to the national level, and this sum was compared with the reported national total. Both the national and sub-national totals were aggregated to yearly scales for comparison with records in the PHAC portal.

For diseases, sub-class totals were aggregated and compared to the overall disease totals. In cases where the sum of sub-diseases was less than the reported disease total, the difference was labeled as disease_unaccounted and included in the normalized data as a derived entry with a record-origin of derived-unaccounted-cases.

For each cross check, we put all records with discrepancies into a CSV file with provenance information for finding the original scans and excel files to more easily fix potential errors. The scripts for producing these CSV files can be found at:

https://github.com/canmod/iidda/tree/main/pipelines/canmod-cross-checks/prep-scripts

A link to download the current state of these CSV files, along with the data themselves, is located at:

https://github.com/canmod/iidda/blob/main/README.md#canmod-digitization-project

We plan to continue addressing these potential issues, but if you use the archive, please check the discrepancy files to see if any data of interest is flagged. Given that our pipelines and tools are open, users are encouraged to fix issues and submit pull requests to our GitHub repository:

https://github.com/canmod/iidda

The following list summarizes the current status of each cross-check:

- Time-scale cross checks

**–** Total number of year-location-disease combinations with discrepancies: 365
**–** Percentage of these combinations without these discrepancies: >99%
**–** Percentage of these discrepancies that are from handwritten data: 100%
- Location cross checks

**–** Total number of period-disease pairs with discrepancies: 2,036
**–** Percentage of these pairs without these discrepancies: >99%
**–** Percentage of these discrepancies that are from handwritten data: 63%
- Disease cross checks

**–** Total number of period-location pairs with discrepancies: 183
**–** Percentage of these pairs without these discrepancies: >99%
**–** Percentage of these discrepancies that are from handwritten data: 88%

It is difficult to combine this information into an estimate of the overall proportion of the harmonized dataset that is error free, because the above estimates are for different stratifications of the data. We can expect errors to yield discrepancies in more than one cross check. For example, a single data-entry error in a sub-disease, province, and week could trigger a discrepancy in all three cross checks if the disease, national, and annual data were reported as well. Not all of these discrepancies represent our data-entry errors because sometimes the original sources are inconsistent or unclear (especially in the handwritten data). Sources also varied in the quality of their marginal totals, and so we removed these totals from our cross checks. But overall, given that the percentages of the combinations of factors that are free of known discrepancies are all greater than 99%, we do not expect that many more data-entry errors remain.

##### Comparing with the PHAC portal

We also compared the national and yearly data on the PHAC portal (https://diseases.canada.ca/notifiable) with aggregated national and yearly totals in CANDID for what we believe are the same diseases. We made these comparisons visually using line plots. Fig H gives an example using chickenpox.

We do not expect the data sources to match exactly, for a variety of reasons. Before 1924 and after 2000 all of the data come from provincial data sources, and so our spatial coverage is limited at these times, whereas the PHAC portal typically reports using data from more provinces after 1924 (though not always, as is evident in the bottom panel of Fig H). CANDID historical source documents presumably included the best numbers at the time. PHAC may have updated these numbers to account for data quality corrections or changes in criteria for determining whether a case qualifies as a specific disease. Discrepancies could also occur because the sub-diseases included in a particular disease change over time.

##### Fixing data

These data quality checks have allowed us to correct errors, and to continue to do so, including data-entry typos (e.g., if 500 cases should have been 50), fixing bugs in preparation scripts, and rethinking the interpretation of data source organization. For instance, the difference between zero incidence and missing incidence was not clear in some of the handwritten data (main text Fig 2). Additionally, we needed to change how we aggregated sub-diseases to be comparable with aggregations on the PHAC portal.

**Fig H:**
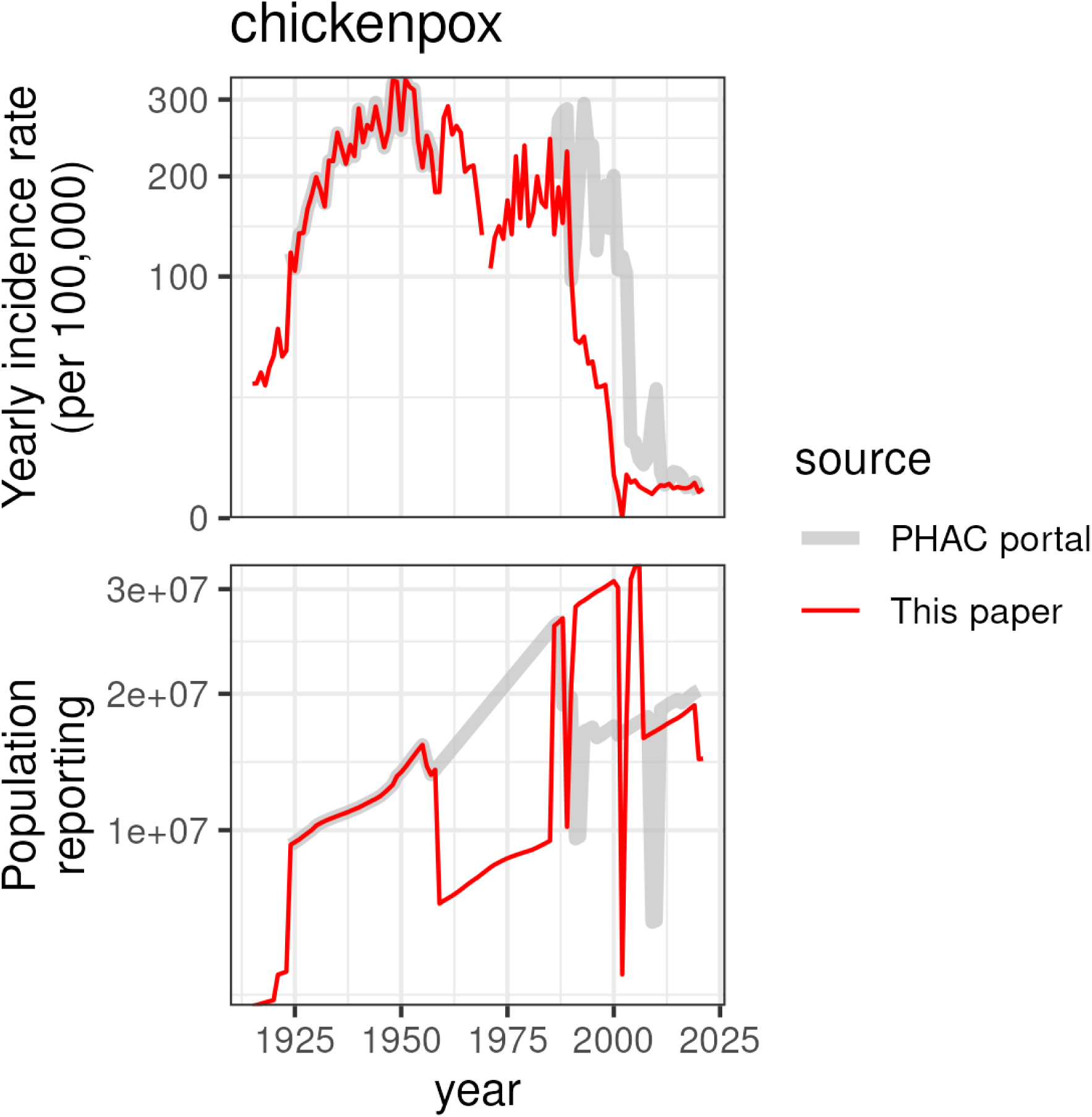
Chickenpox as an example of a comparison between our archive and the PHAC portal. The top panel gives yearly incidence from CANDID in red and the PHAC portal in grey. The bottom panel gives the total population of all reporting provinces for each source. Jumps in this bottom panel are caused by provinces being added or dropped from each source. For chickenpox the reported incidence is very similar in both sources during times when the reporting population is identical. Comparisons for all diseases that are shared by both datasets can be found on the website for the paper: https://github.com/canmod/candid/tree/main/output/phac-portal-comparisons.

#### J Polio methods

We analyzed weekly provincial polio data. For each province and week, we calculated the weekly incidence rate by multiplying the number of new cases by 100,000 and dividing by the interpolated population for that week. To estimate the national incidence rate, we summed the provincial cases and divided by the population of provinces with available data, then multiplied by 100,000. Vertical lines in the plot (main text Fig 7) indicate the week with the highest national incidence in a given polio year, which we defined as the 52-week period centered around epidemiological week 34, the typical yearly peak for polio. We plotted peaks only for years with more than 20 total cases reported in the country.

We identified the peak polio week for each combination of province and polio year. We defined the peak as the week in the polio year with the highest incidence. To ensure sufficient data for estimating a clear peak, we excluded combinations with sparse or limited case counts (≤ 5 weeks of non-zero incidence or ≤ 20 total cases). Fig I shows the distribution of peak weeks by province and highlights that most provinces in most years peaked between August and October.

#### H Whooping cough methods

We estimated the annual incidence rate in each of the six geographic regions of Canada, and Canada as a whole, as follows. We first computed the average daily number of whooping cough cases for each province or territory by summing the reported cases over all available time periods within a year and dividing by the total number of observed days. We then estimated the total annual cases for each province by scaling this daily rate by the total number of days in the year, accounting for any missing data. Summing these estimates across the provinces within each geographical region gave us the total estimated cases per region. Finally, we calculated the incidence rate per 100,000 individuals by dividing the regional total estimated cases by the total regional population and scaling appropriately. These sums and averages were not contaminated by double counting, as each reported or implied case is represented only once in the normalized dataset (see the section on Preparing the normalized CSV file). However, our method could be affected by within-year variation in incidence rates that did not average out over the sample of available time periods.

Details for these calculations are as follows:

- Let *x_ij_* be the number of new whooping cough cases reported during time period *i* (e.g., week, month, or quarter) within province or territory *j* of Canada.
- Let *n_ij_* be the number of days in time period *i* within province or territory *j*.
- Let Ω*_k_* be the set of all time periods *i* within year *k*. Note that Ω*_k_* may not include all possible periods if data are missing for some weeks, months, or quarters.
- Let Ψ*_l_* be the set of provinces and territories *j* that are contained within geographical region *l* of Canada (e.g., Atlantic, Quebec, Ontario, Prairies, British Columbia, Territories).
- Let *N_k_* be the total number of days in year *k* (either 365 or 366).
- Let *p_kj_* be the population of province or territory *j* during year *k*.

**Fig I:**
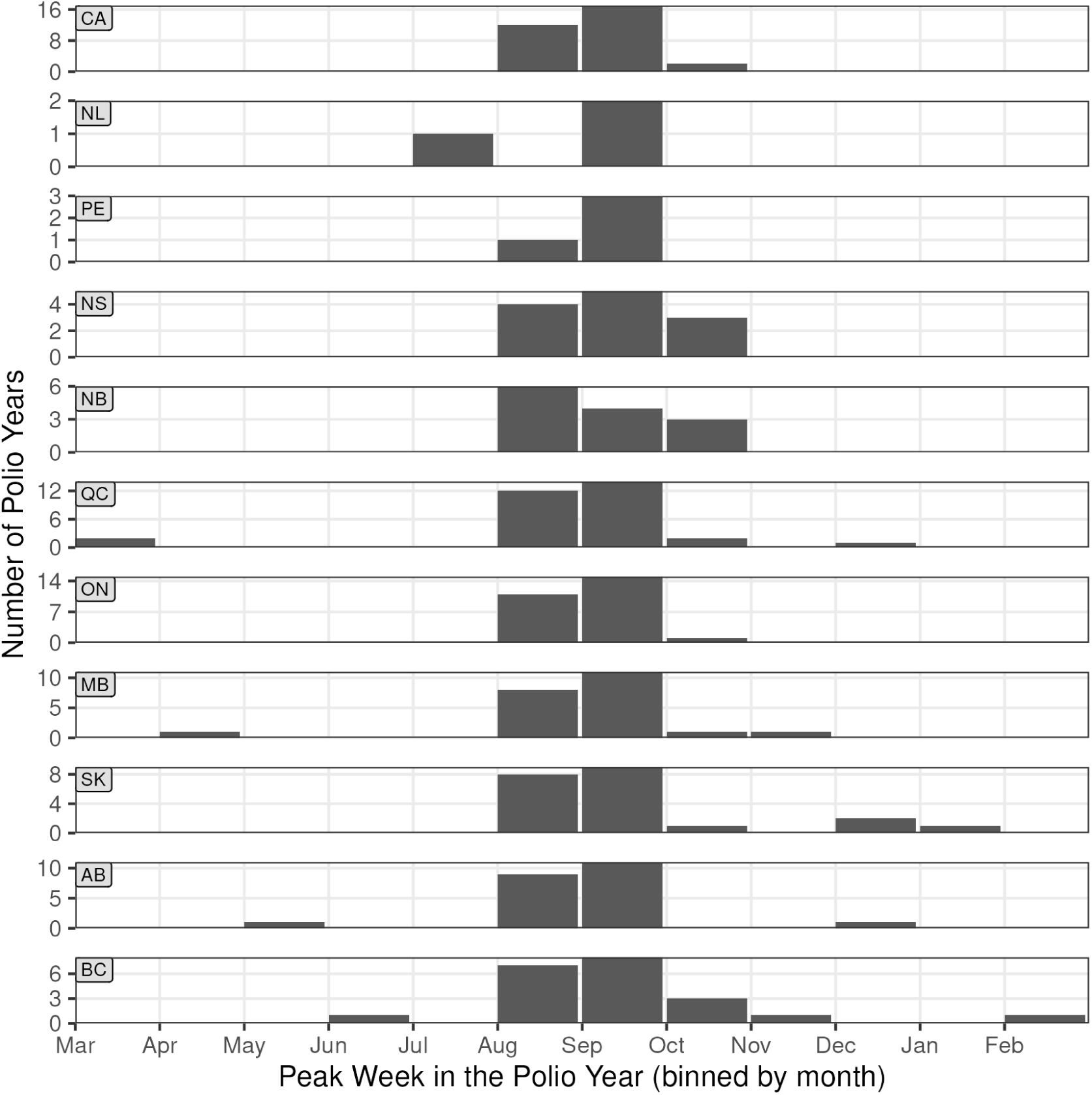
Histograms of the distribution of the peak week binned by month. Each panel gives the distribution for Canada (top panel) and each province.

1. Average Daily Cases in Year *k* and Province *j*:

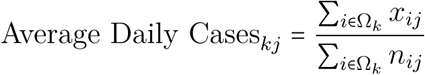
2. Estimated Total Annual Cases in year *k* in Province *j*:

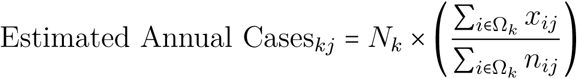
3. Total Estimated Cases in Year *k* and Region *l*: Sum the estimated annual cases over all provinces and territories *j* within region *l*:

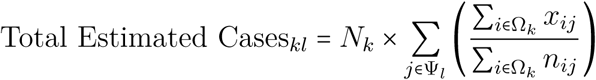
4. Incidence Rate per 100,000 Individuals in Year *k* and Region *l*: Calculate the incidence rate by dividing the total estimated cases by the total population of the region and scaling by 100,000:

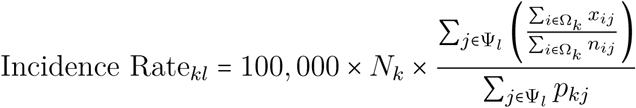

## Footnotes

1 Typically these strings were taken as is from the source, but in many instances before 1924 we had to make an educated guess about the reason why particular records were missing.

